# Learning Patient Similarity from Genomics for Precision Oncology

**DOI:** 10.64898/2025.12.17.25342480

**Authors:** Maha Shady, Brendan Reardon, Sharon Jiang, Erica Pimenta, Tess O’Meara, Jihye Park, Kenneth L. Kehl, Haitham A. Elmarakeby, Shamil R. Sunyaev, Eliezer M. Van Allen

## Abstract

**Introduction:** Precision oncology has informed cancer care by enabling the discovery and application of diagnostic, prognostic, and/or predictive molecular biomarkers. However, many patients lack actionable biomarkers or fail to respond to biomarker-directed therapies. Patient similarity approaches can leverage comprehensive tumor profiling and prior clinical experiences from large cohorts for decision support, facilitating broader realization of precision oncology insights.

**Methods:** We developed a deep learning-based modeling framework using real-world clinicogenomic data from a tertiary cancer center to (i) measure patient similarity based on embedded tumor genomic profiles and (ii) evaluate the association of derived patient subgroups and neighborhoods with shared therapeutic outcomes in breast cancer-specific and histology-agnostic pan-cancer settings.

**Results:** The model recovered clinically meaningful patient clusters reflecting both expected and previously unknown therapeutic associations, as well as patient-specific neighborhoods that could inform therapeutic trajectories more often than expected by chance in multiple clinical contexts. Moreover, model utility extended to patients without actionable genomic biomarkers and those with cancer of unknown primary (CUP) diagnoses, where neighborhoods aligned with independently predicted primary cancer type. These neighborhoods could also be examined over time in a continuously learning scenario.

**Conclusion:** This similarity-based modeling framework distilled complex molecular and clinical data into concise, context-specific insights that augment clinician judgment, providing a foundation for a real-time learning, patient-centered decision support model in precision oncology.

## Introduction

Precision oncology, which constitutes the use of an individual patient’s tumor molecular profile and clinical context for diagnostic, prognostic, and/or therapeutically predictive purposes, has transformed cancer patient care and improved outcomes for many patients^1–3^. These personalized treatment decisions are enabled by advances in next-generation sequencing technologies that have made comprehensive genomic profiling of tumors more ubiquitous in clinical settings^4,5^. There are now numerous regulatory approvals and clinical guidelines that indicate molecularly guided treatments for patients with cancer based on specific genomic alterations or representative genomic features, such as microsatellite instability (MSI) or tumor mutational burden (TMB)^6–9^. While many of these indications are histology specific, there has been an increase in multi-histology or histology-agnostic approvals^9–14^. Even with continued improvements in clinical outcomes, many patients still have limited or no response to their matched therapies, despite eligibility, and many others have tumors that are negative for clinically actionable genomic features^15,16^. A more comprehensive view of a tumor’s genomic profile may help uncover specific alterations, genetic interactions, or high level genomic features that contribute to diagnostic factors or selective response to therapeutic agents, expanding the benefit of precision oncology guided strategies.

Recent studies have aimed to improve drug recommendations and response prediction by taking advantage of the expanding molecular and clinical data corpus for patients with cancer. Prospective clinical trials have utilized comprehensive genome and transcriptome sequencing of tumor samples to match patients to single-agent or multi-agent therapy regimens based on known therapeutic targets and biological associations^17,18^. Others have leveraged real-world clinicogenomic datasets to identify potential therapeutic or prognostic biomarkers and develop clinical decision support systems^19^. Such studies have identified potential co-mutations or clinical-marker interactions with prognostic or therapeutic outcome prediction value^19–23^, as well as emulated randomized controlled trials to identify patient cohorts likely to benefit from trial outcomes^15^. However, the ability to integrate molecular profiles and prior patient clinical trajectories into a patient-specific context for generalized precision oncology decision support remains incompletely realized.

To begin addressing these challenges, the concepts of ‘virtual patients’ and ‘digital twins’ have been increasingly evaluated in oncology research and clinical translation^24–28^. ‘Virtual patient’ studies entail simulating a patient population from mechanistic or statistical models and can enable clinical trial simulation or population-level hypothesis generation, while ‘digital twin’ studies involve generating *in silico* representations of an individual patient that can be continuously updated along with the patient’s disease trajectory and utilized to optimize treatment strategies for that patient. Recent studies have demonstrated the potential of these approaches to aid oncology treatment selection in specific disease contexts^25,29–32^. While promising, these studies have been limited in scope (e.g. often lacking inclusion of molecular data and focusing on restricted patient cohorts or clinical scenarios). Moreover, the development of generalized frameworks incorporating digital twins that can be used in clinical settings will require extensive data generation and integration, as well as mechanistic model development that can enable simulation of plausible outcomes^24–28^.

The concept of ‘patient similarity’ is another recurring theme in precision medicine studies as it may provide a comprehensive, patient-centered approach to personalize clinical care^33–37^. Unlike digital twin studies, patient similarity approaches rely on a broad suite of prior patient experiences and do not require simulation of hypothetical patient scenarios. To date, similarity applications in oncology have focused on tumor profile to cell line matching^1,38^, overall risk or subtype prediction^39–42^, or applications in narrowly defined patient subgroups^41,43,44^. These prior works indicate the potential of patient-similarity based frameworks in precision oncology (especially for patients who lack actionable genomic biomarkers), but are limited in scope and methodologic development. Critically, the increasing availability of integrated molecular and clinical data of patients with cancer, along with advances in artificial intelligence (AI)-guided modeling, pave the way for generalized cancer patient similarity models that may aid clinical decision making across a wide range of precision oncology clinical scenarios and reduce the need for numerous task-specific models.

Here, we developed a deep learning framework using real-world clinicogenomic data from a tertiary cancer center^5,45^ to model patient-to-patient tumor genomic similarity and evaluate associations with shared therapeutic outcomes. We show that genomic similarity corresponds to therapeutic response similarity in both a breast cancer-specific and a histology agnostic pan-cancer model. We further demonstrate the framework’s potential utility in various clinical contexts, including as a diagnostic aid for cancer of unknown primary (CUP) cases, and as a versatile learning system that adapts with evolving clinical paradigms. Our approach enables data-driven clinical decision support by providing clinicians with therapeutic trajectories of the most genomically similar prior patients to the patient being treated. Thus, our framework is designed to support clinician decision-making rather than produce prescriptive predictions, thereby maintaining clinician agency and enabling broadly applicable decision support. This first demonstration of a generalized cancer patient genomic similarity model with clinically relevant representations provides a template for continuous learning implementations across cancer centers.

## Results

### Similarity Framework

We established a methodologic framework for determining tumor genomic similarity between patients and assessing its association with longitudinal clinical oncology outcomes (Methods; Figure 1; Supplementary Figure 1). Briefly, patient tumor similarity was determined based on learned embeddings of the tumor genomic profiles (via the OncoPanel hybrid-capture sequencing assay^5^) at the time of sequencing. After evaluation of multiple modeling strategies, genomic embeddings were created using an autoencoder model trained on features extracted from the assay results (Supplementary Table 1). To evaluate the clinical relevance of these genomic embeddings, we separately utilized electronic health record (EHR) data that includes medications administered as well as medical notes and radiology reports to infer patient-specific therapeutic trajectories^45^. Groups of genomically-similar samples based on the learned embeddings were analyzed at two levels of granularity: (i) K-means clustering was applied to the genomic embeddings to define groups of genomically similar patients and then evaluated based on shared therapeutic trends, and (ii) K-nearest neighbors analysis was used to define patient-specific neighborhoods based on genomic embeddings, which were evaluated for the potential to highlight personalized therapy recommendations at the point of care based on the retrieved therapeutic trajectories. This was done for two cohorts: (i) a breast cancer cohort (“breast cancer model”; n = 3,496 patients), and (ii) a combined cohort of solid tumors (“pan-cancer model”; n = 27,577 patients) (Methods; Supplementary Table 2).

**Figure 1:**
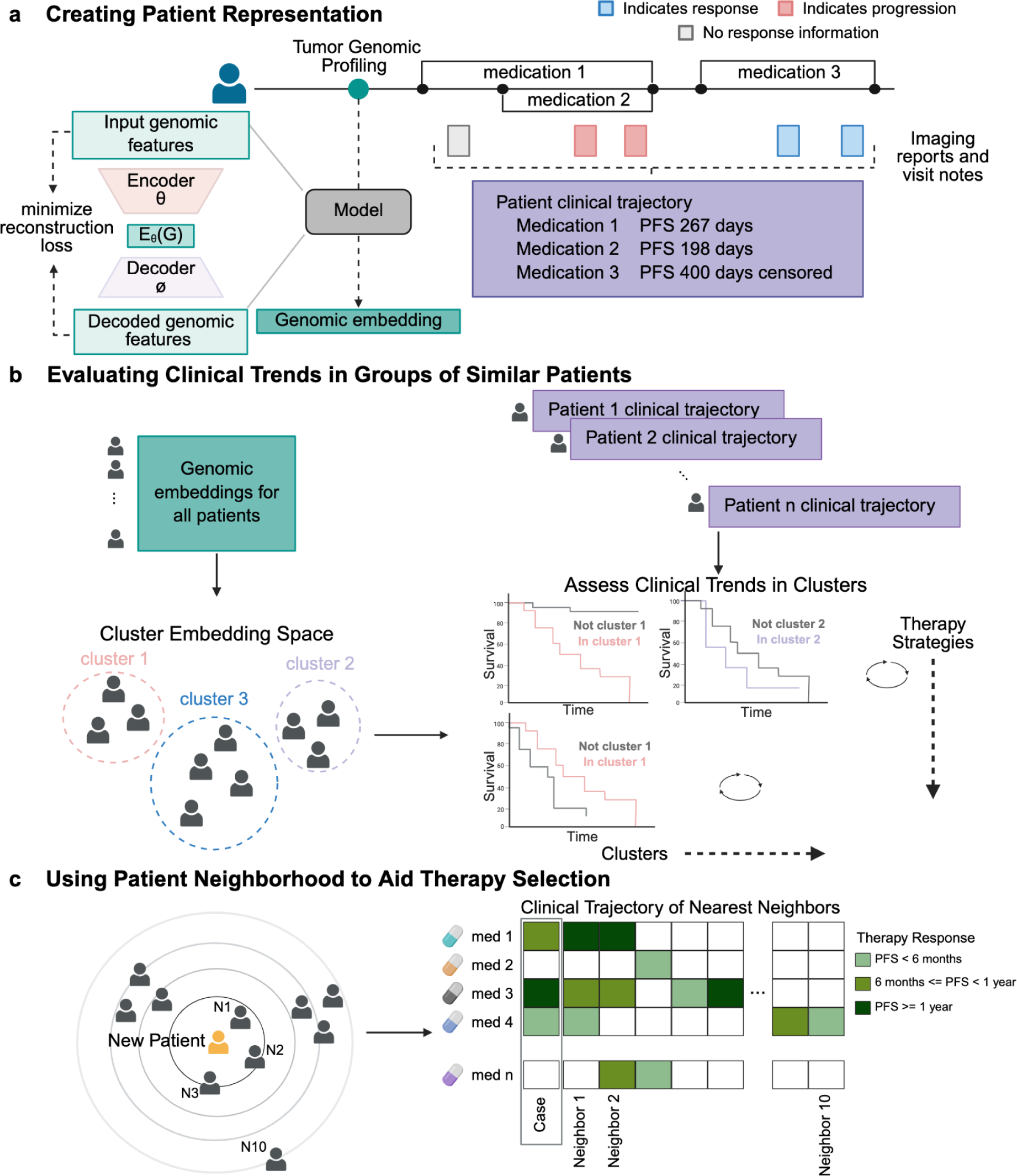
Framework overview. a: Somatic genomic alterations from patient tumor samples were used as input for an autoencoder model to create genomic embeddings. Post-sequencing clinical events included medications administered, and clinical and imaging reports. These data were processed into clinical trajectories of progression free survival (PFS) for each medication class per patient. b: Genomic embeddings from all patients were clustered, and clusters were interpreted based on their defining genomic biomarkers as well as whether they separated patients by response to specific therapy types (Methods). c: The nearest neighbors to each patient were evaluated for their relevance, defined by whether they shared the PFS category on administered therapy types with the case patient. This was summarized into an average precision score for each patient’s neighborhood, and as a percentage of neighborhoods defined as useful (average precision at 10 neighbors >= 0.2; Methods; Supplementary Figure 1) for the whole cohort.

### Breast Cancer Patient Similarity Model Reveals Known and Previously Unknown Therapeutic Trends

We first trained our genomic autoencoder and clustering framework on 2,231 tumor samples from 2,097 patients with breast cancer and applied it to a held-out validation set of 1,480 samples from 1,399 patients (n = 3,496 total breast cancer patients). The framework derived clusters that separated samples by genomic features and therapeutic trajectory (Figure 2).

**Figure 2:**
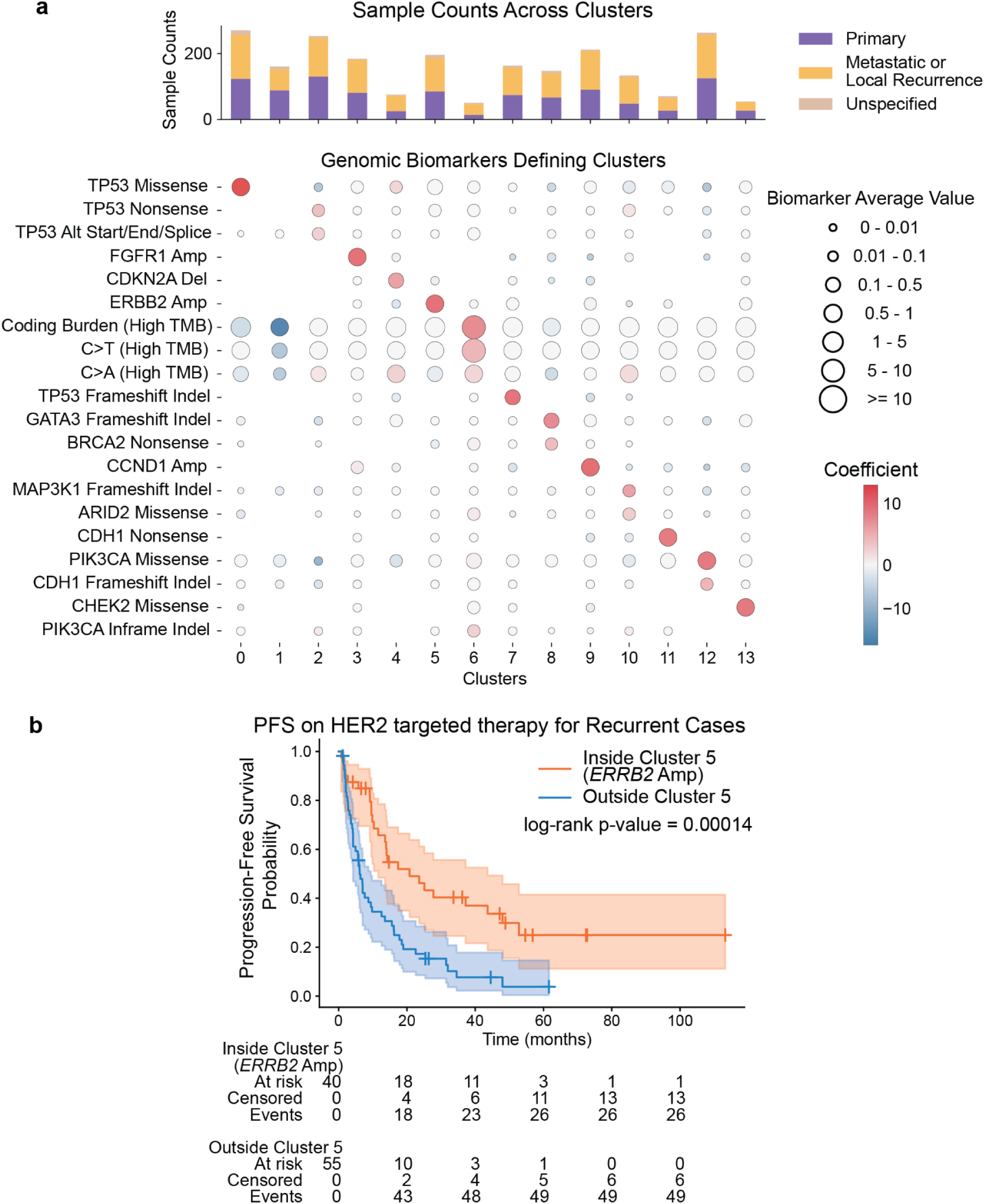
Breast Cancer Model Clusters. a: Genomic features enriched within each cluster. Features included are those with a coefficient of at least 2.5 for any of the cluster membership predictive models and an average value of at least 0.1 in the corresponding cluster. Circle color indicates coefficient for a given cluster, with blue indicating a negative coefficient implying a negative association, and red indicating a positive coefficient. Circle size indicates the average value of the feature among training set samples in each cluster. 7/14 clusters (clusters 0, 3, 5, 7, 9, 11, and 13) were enriched for a single mutation, while the others were defined by a combination of features rather than a single mutation. The bar plot shows the number of primary and recurrent/metastatic samples in each cluster. b: Trend of improved PFS on HER2 targeted therapy for samples in the *ERBB2* amplification defined cluster (cluster 5) based on biopsies taken from locally recurrent or metastatic tumors.

To interpret cluster membership, we separately trained regularized logistic regression models to predict each cluster’s membership based on unembedded genomic features (Methods). Models trained to predict embedding-derived cluster membership based on unembedded genomic features exhibited high performance on both the training and validation sets (training AUROC: 0.982-1, AUPRC: 0.852-1; validation AUROC: 0.912-0.999, AUPRC: 0.569-0.985; Methods), suggesting that the autoencoder and clustering framework was robust and generalizable (Supplementary Table 3). Half (7/14) of the clusters identified were enriched for a mutation type in a particular gene, including expected breast cancer clusters (Figure 2a; Methods). For example, cluster 0 was enriched for samples with *TP53* missense mutations, cluster 5 for *ERBB2* amplified samples, and cluster 9 for *CCND1* amplified samples. Certain clusters were defined by combinations of features, rather than associating with a singular somatic mutation. For instance, cluster 8 membership was reliably predicted with the trained logistic regression model (training AUROC = 0.984, AUPRC = 0.887; validation AUROC = 0.968, AUPRC = 0.776) using a combination of somatic events; most strongly *GATA3* frameshift indel variants along with *BRCA2* nonsense mutations, among other somatic features (Supplementary Table 4). Similarly, membership of cluster 12 could be predicted by a combination of features that include *PIK3CA* missense mutations and *CDH1* frameshift indel variants, among others (Methods; Supplementary Table 4). We additionally identified low and high mutational burden clusters (clusters 1 and 6, respectively; Methods).

We next assessed cluster-specific therapeutic trends in the subset of patients with therapeutic labels (n = 993 and 662 patients in training and validation sets, respectively). We first investigated cluster 5, which was enriched for samples carrying somatic *ERBB2* amplifications. As expected, this cluster was enriched for patients receiving HER2 directed therapy (59/144 and 39/102 of HER2 directed therapy instances in training and validation sets respectively; Supplementary Table 5), and membership of cluster 5 was associated with improved progression-free survival (PFS) on HER2-directed therapy for metastatic or locally recurrent cases^8,46,47^(Figure 2b; training log-rank test p-value = 0.00014, validation p-value = 0.0534; Supplementary Table 6). This observation was a reassuring indication that the model recapitulated an expected, well validated biomarker association^8,46^.

In certain cases, clusters suggested potentially relevant associations that have some evidence in literature but were not definitive clinical guidelines. For example, we observed reduced PFS following treatment with endocrine therapy (aromatase inhibitors and ER signaling inhibitors) for samples in cluster 9 (training log-rank p-value = 0.00925, validation p-value = 0.00132; Supplementary Table 6), which was defined by *CCND1* amplifications (Figure 2a; Supplementary Figure 2a). A subset analysis was performed on biopsy samples taken from primary breast tumors, which showed that cluster 9 membership was only associated with shorter PFS on endocrine therapy either in the primary or metastatic setting when patients had received at least 1 prior line of endocrine therapy and/or CDK4/6 inhibition (training log-rank p-value = 0.0673, validation p-value = 0.0801). There was no association between cluster 9 membership and shorter PFS on endocrine therapy when it was the first line of endocrine therapy and/or CDK4/6 inhibition received. Previous studies have reported an association between *CCND1* amplification and poor prognosis^48^, as well as reduced response to aromatase inhibitors in breast cancer^49,50^. While these studies particularly considered patients who presented with metastatic breast cancer and have received no prior breast-cancer related therapy, the patient similarity model primarily observed a trend among patients who received prior lines of therapy. Furthermore, we observed a set of *ERBB2/CCND1* co-amplified samples in cluster 9 and found that these patients had reduced PFS on HER2 directed therapy compared to all other patients with *ERBB2* amplified samples in the training set (logrank p-value = 0.0011; only 4 samples in the validation set received HER2 therapy, which did not satisfy our minimum thresholds for running subset analyses (Methods)). Indeed, a similar trend of reduced PFS on HER2 directed therapy in cases with *CCND1* amplification was recently reported^51^. These results suggest the model can refine risk stratification beyond single-gene associations.

Building upon therapeutic trends that recapitulated standard clinical practice or findings previously suggested in literature, we also identified additional previously unexplained trends that may prompt additional investigation. For instance, we observed improved PFS on CDK4/6 inhibitors (training log-rank p-value = 0.023, validation p-value = 0.0591) for cases in cluster 1, defined by low tumor mutational burden (TMB) (Figure 2a; Supplementary Figure 2b; Supplementary Table 6). Whether this relationship is causally linked to low TMB or some correlated feature (e.g. low tumor purity due to increased immune infiltration, among others) is unknown.

To further assess the overall reliability of the clinical trends detected by the model, we ran the Cauchy combination test^52^ on all the log-rank test results, which provides a combined p-value and does not assume independence of individual tests. Our model yielded a combined p-value of 0.0059 on the training set and 0.005 on the validation set. Upon removing the expected anti-HER2 therapy trend in cluster 5, the similarity model remained significant (training combined p-value of 0.0071 and a validation combined p-value 0.0049). These results suggest that at least one of the trends identified by the model, other than the HER2 association, was statistically significant. Finally, to complement this assessment of clinical structure identified by the model, permutation testing also indicated a low probability that randomly sampled groups would define a structure as clinically meaningful as the proposed framework (p-value = 0.00014; Methods).

### Using Patient Similarity Model to Aid Individualized Treatment Decision-Making in Breast Cancer

Given the model performance for aggregate patient similarity clustering, we next evaluated how an individual patient’s 10-nearest neighbors in the learned genomic embedding space could be used for hypothetical clinical decision support at the point of care (Methods). As a case study, we selected a *ERBB2/CCND1* co-amplified case from cluster 9 that had a PFS of less than 6 months on HER2-directed therapy to illustrate how the case’s clinical trajectory could have been informed by its neighborhood (Figure 3a). This case was a post-menopausal woman diagnosed with stage 3 breast cancer, ER+ PR+ HER2 3+, who received neoadjuvant chemotherapy, breast surgery, 1 year of adjuvant trastuzumab, and 6 years of adjuvant endocrine therapy (tamoxifen followed by aromatase inhibition). She then experienced a metastatic recurrence 6 years after initial diagnosis, with confirmed ER+ PR+ HER2 3+ osseous metastatic disease. Genomic sequencing on this metastatic bone biopsy showed *ERBB2*/*CCND1* co-amplification and was used to construct nearest neighbors. The patient then very quickly progressed through multiple lines of HER2-directed therapy. Nearest neighbors to this patient also demonstrated short time on both endocrine therapy and HER2-directed therapies. Equipping the treating oncologist with this information upfront may have justified alternative treatment options involving clinical trials or other escalated therapeutic strategies (e.g. upfront trastuzumab deruxtecan (T-DXd)) for this patient, given the likelihood of progression based on the neighborhood.

**Figure 3:**
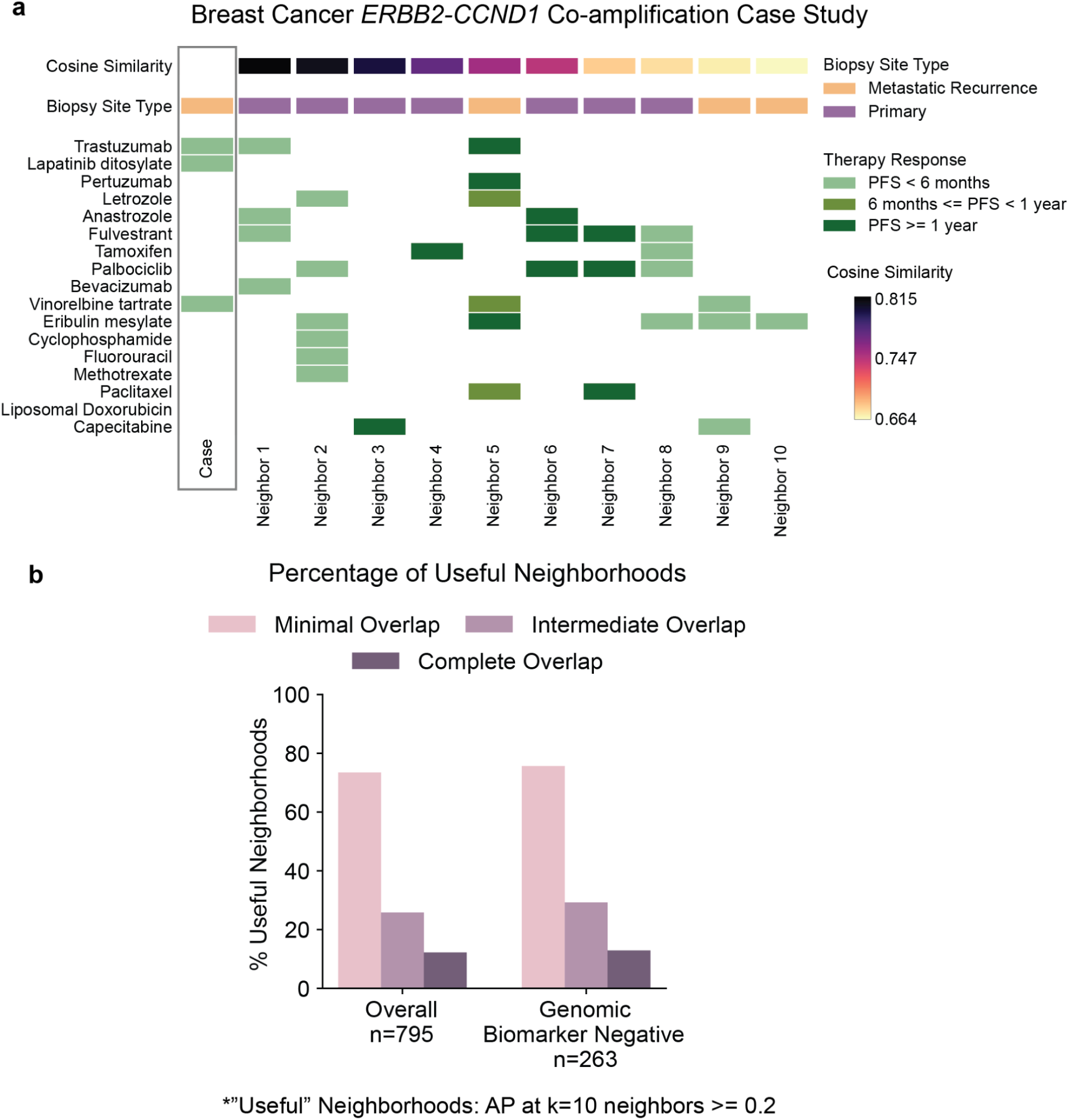
Breast Cancer Model Neighborhoods. a: A case with *ERBB2/CCND1* co-amplification. Therapeutic trajectories of nearest neighbors show reduced response to HER2-directed and endocrine therapies (neighbors 1, 2, and 8), and may suggest that the target patient may also not respond well to those therapeutic strategies, as was indeed observed retrospectively for HER2 directed therapy. b: Percentage of patients with useful (average precision value at k=10 neighbors >= 0.2) neighborhoods in the full breast cancer training cohort, as well as the genomic biomarker-negative sub-cohort (Methods). Bar colors indicate the level of therapeutic overlap required to consider a neighbor as relevant (Methods). Retrospective precision estimates were used for neighborhood utility categorization due to the need to consider therapies administered to the target case. Proportions of useful neighborhoods with variable stringency levels illustrate how often a new case may yield clinically informative neighborhoods.

To broadly assess neighborhood utility and obtain a cohort-wide metric, we calculated the average precision at the 10th neighbor for each sample in the cohort (“AP at k=10”; Methods) and computed the percentage of samples whose neighborhood AP at k=10 values were at least 0.2 (categorized as “useful” neighborhoods; this threshold was chosen based on performance benchmarking of tumor-to-cell line matching models^1^). To account for different clinical scenarios, we performed these analyses across varying levels of stringency: (i) “complete” overlap, requiring identical clinical trajectory for a neighbor to be considered relevant; (ii) “intermediate” overlap, requiring all of the case’s labels being shared by the neighbor; or (iii) “minimal” overlap, having at least one shared therapy administered with the same corresponding outcome as sufficient for neighbor relevance (Methods, Supplementary Figure 1). This tiered approach was taken since partial overlap may still provide clinically meaningful information. This approach also addresses the practical limitation that complete overlap cannot always be assessed, as patients may not have received every comparable therapy, and provides clinicians with flexible thresholding options to use at their discretion. We note that the average precision values were retrospectively computed based on the therapies received by the target case and, therefore, would not be available in a prospective setting. However, the proportion of cases with reported “useful” neighborhoods can serve as an estimate of how likely it is that a new case would have a similarly informative neighborhood.

For the case study presented, AP at k=10 neighbors was 0.664 based on minimal therapeutic overlap, and 0 based on both intermediate and complete therapeutic overlap. This was expected because none of the nearest 10 neighbors received exactly the same medications as the target case, rendering utility based on intermediate and complete therapeutic overlap unevaluable in this case (Methods). We then extended our analysis to the full training cohort of breast cancer samples. We observed that 73.5%, 25.8%, and 12.2% of cases had useful neighborhoods when considering minimal, intermediate, or complete therapeutic overlap, respectively (Figure 3b). Permutation testing, in which each case was assigned a random set of 10 neighbors, showed that these percentages were above what is expected by chance (p-values = 0.0002 for each level of overlap). To assess the generalizability of our results to unseen patients, we identified the nearest neighbors among the training set samples for each case in the validation set (n=522) and computed the percentage of validation set cases with useful neighborhoods. We found that 74.9%, 21.3%, and 8.4% of cases had useful neighborhoods under minimal, intermediate, and complete therapeutic overlap, respectively.

To assess whether these values were driven primarily by cases carrying genomic biomarkers associated with FDA-approvals or clinical guidelines in breast cancer, we removed all cases carrying such biomarkers (Methods; Supplementary Table 7) and recomputed the percentage of samples with useful neighborhoods (AP at k=10 neighbors >= 0.2) for the remaining 263 samples. Among the genomic biomarker negative group (n=263), 75.7%, 29.3%, and 12.9% of samples had useful neighborhoods with minimal, intermediate, and complete therapeutic overlap for neighbor relevance, respectively (Figure 3b; p-values = 0.0028, 0.0004, 0.002, respectively). These results, along with the case study presented, reveal the potential utility for this framework as a decision-making aid for biomarker negative cases, as well as for additional risk stratification in genomic biomarker positive cases.

### Pan-cancer Patient Similarity Model Expands Utility and Reveals Cancer-Type Agnostic Patterns

Given the performance of the single tumor type patient similarity model, we next applied the machine learning framework to a larger set of solid tumors (Methods). Our pan-cancer training cohort consisted of 17,615 sequenced samples from 16,522 patients, with therapeutic labels associated with 6,097 samples from 5,705 patients. The validation cohort consisted of 11,804 samples from 11,055 patients with therapeutic labels associated with 4,026 samples from 3,723 patients (total n = 27,577 patients; Supplementary Table 2).

This approach resulted in 24 pan-cancer clusters representing the genomic embedding of this cohort (Figure 4a). All clusters included samples from more than one cancer type, and only 4 out of 24 (16.67%) clusters were majority populated by a single cancer type (>50% of cluster members belong to that cancer type; Supplementary Table 8). As expected, multiple clusters primarily grouped cancer types known to share the cluster-defining biomarker (Methods; Supplementary Tables 8, 9). For example the *BRAF* missense mutation cluster (cluster 18) was enriched for samples from the melanoma, thyroid, colorectal, and non-small cell lung cancer cohorts, and the *KRAS* missense mutation cluster (cluster 23) was enriched for pancreatic cancer, non-small cell lung cancer, and colorectal cancer samples^4,53^ (Figure 4a, b). The pan-cancer model also identified clusters that were predicted by a combination of genomic features even if those features were only present in a subset of the samples in the cluster (Figure 4b; training AUC >= 0.95, AUPRC >= 0.173; validation AUC >= 0.935, AUPRC >= 0.218; Methods; Supplementary Table 3). Examples include cluster 13, which was associated with a feature set that included *PIK3CA* missense mutations and *CDH1* frameshift indel mutations; cluster 14, associated with a set that includes *SMAD4* and *TP53* missense mutations; cluster 17, associated most strongly with *APC* mutations, along with others like *TP53* and *FBXW7* mutations; and cluster 12, which was enriched for high TMB and mismatch repair deficient (dMMR) samples (37% and 31% of dMMR samples in this cluster in training and validation sets, respectively) (Supplementary Table 9). Additionally, most clusters included varying amounts of cancer of unknown primary (CUP) samples (1.49% - 10.1% of cluster members are CUP samples; Supplementary Table 8).

**Figure 4:**
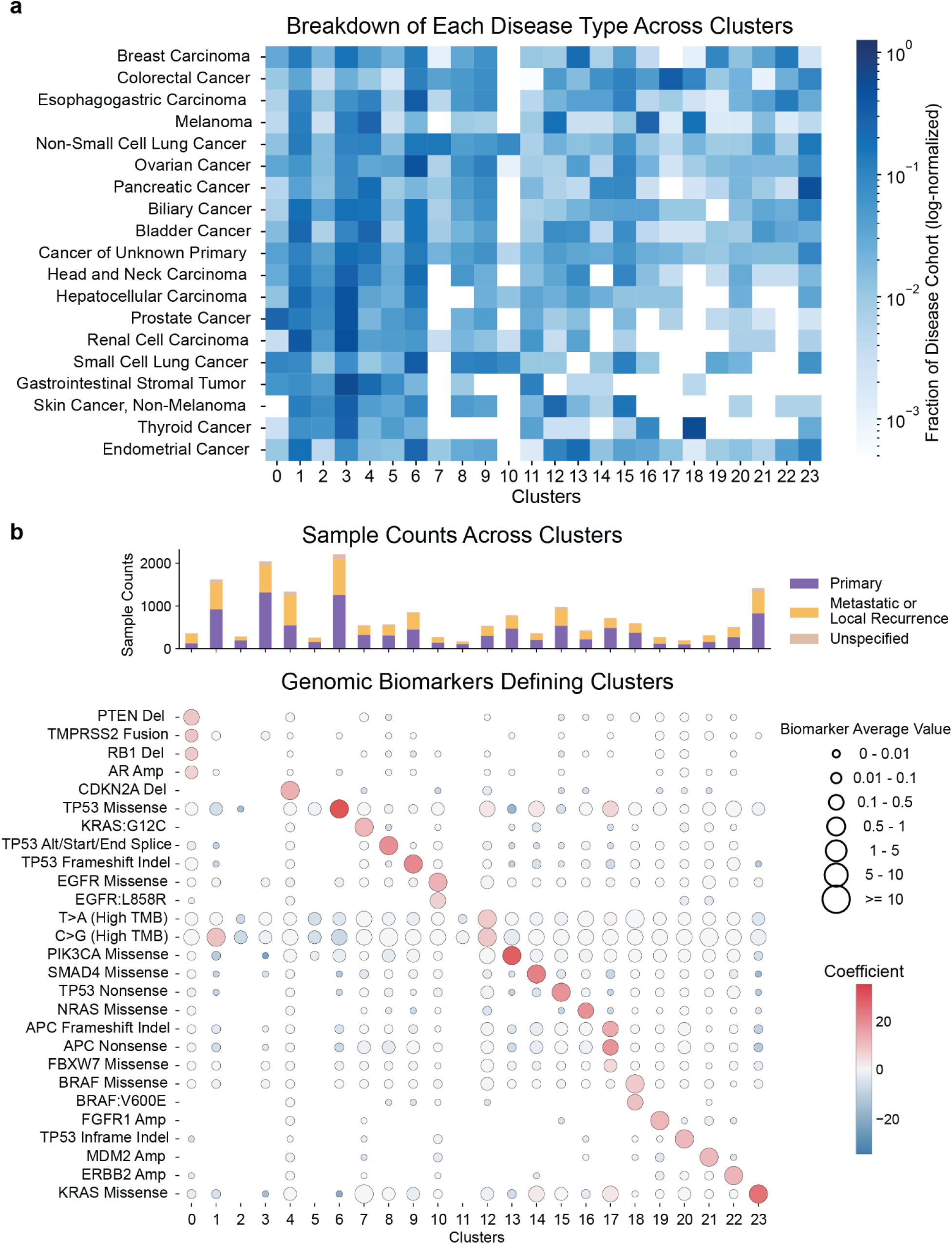
Pan-cancer Model Clusters a: Heatmap of the fraction of each disease cohort assigned to each cluster (log-normalized). b: Genomic features enriched within each cluster. Features included are those with a coefficient of at least 5 for any of the cluster membership predictive models and average value at least 0.2 in that cluster, circle color indicates coefficient of the feature in the predictive model inference for a given cluster and circle size indicates average value of the feature in each cluster. Clusters 0, 1, 2, 3, 5, 11, 12, 13, 14, and 17 are defined by a combination of features rather than being strongly associated with a single mutation. The bar plot shows the number of primary and recurrent/metastatic samples in each cluster.

The pan-cancer model identified both cancer type specific and agnostic trends (Figure 5; Supplementary Figures 3, 4, 5). For example, cluster 12 (high TMB and dMMR samples) patients had improved PFS on cytotoxic chemotherapy (platinum-based training p-value = 4.3×10^-10, validation p-value = 3.44×10^-5; non-platinum based training p-value = 3.04×10^-13, validation p-value = 4.59×10^-8), and PD-1/PD-L1 inhibitors (training p-value = 8.58×10^-12, validation p-value = 2.07×10^-3), even when limiting to non-curative intent treatment instances (Supplementary Figures 4, 5; Supplementary Table 6). When stratified by cancer type, we found that the PD-1/PD-L1 inhibition signal was driven by the melanoma, colorectal, and non-small cell lung cancer cohorts; whereas the chemotherapy signal was driven by colorectal, endometrial, and non-small cell lung cancer samples. This signal was expected due to the cluster enrichment for high TMB and dMMR samples^6,54^. We note that the pan-cancer trends on these therapy classes remained even if we removed samples from the cancer types mentioned, implying that the trends were disease-agnostic (Supplementary Figure 6). This finding further suggests that the pan-cancer model may increase power to detect therapeutic associations across multiple cancer types, including those that may be missed when analyzing each cancer type in isolation.

**Figure 5:**
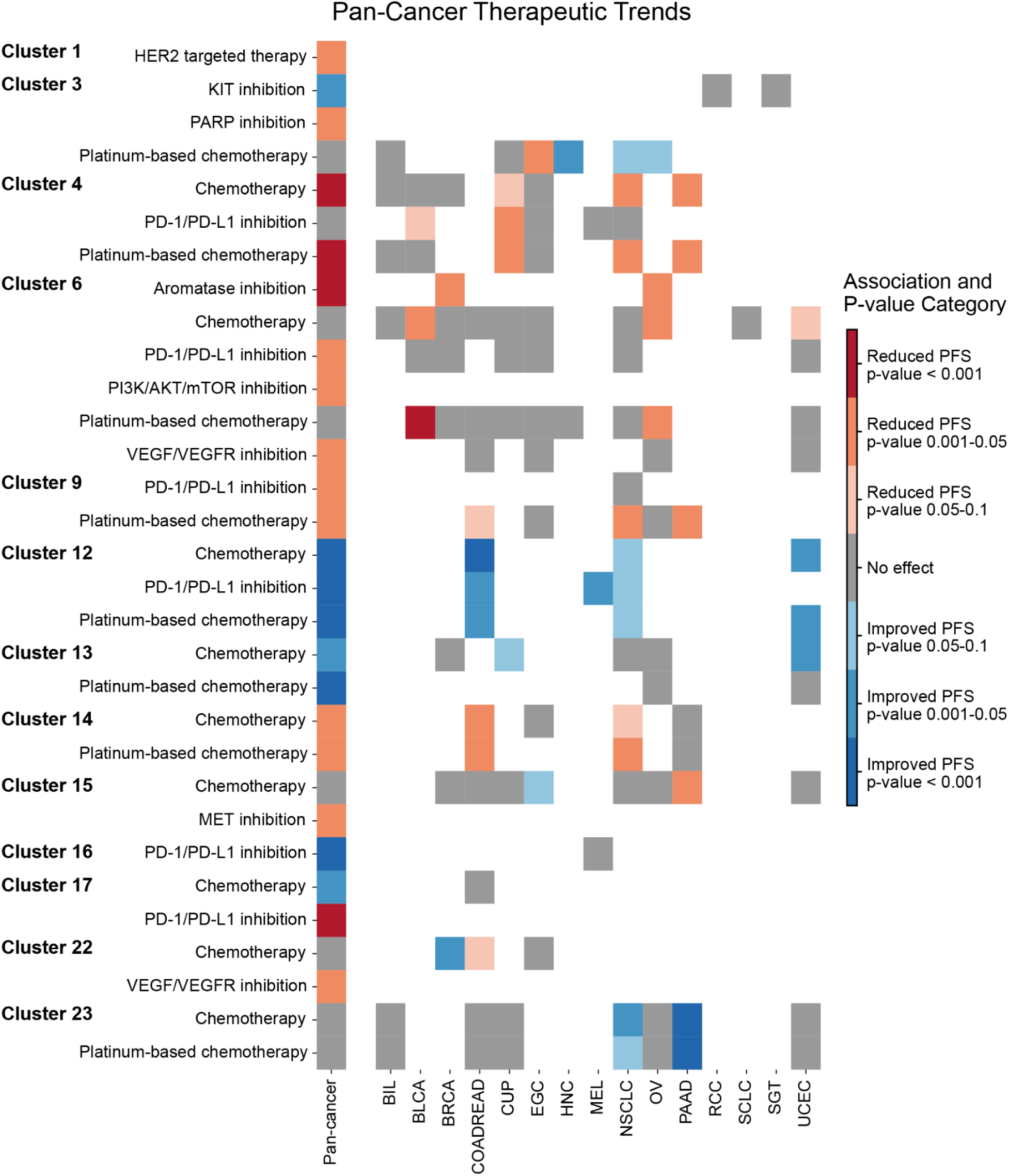
Pan-cancer Model Clinical Trends. Cluster-specific clinical trends with significant or near-significant “pan-cancer” survival differences as assessed by log-rank test. Each row is limited to a specific therapy type and considers patients inside vs outside a specific cluster who received that therapy. Each column represents a cohort: the first column is the pan-cancer cohort and the subsequent columns are limited to samples associated with a specific primary diagnosis as listed in the x-axis labels. Specific cancer types included here are the ones with a significant test for at least one cluster-therapy combination included in the plot. The trends shown either appear in more than one cancer type, or are only evaluable/significant in the pan-cancer setting and not in any one cancer type on its own (e.g. cluster 17 with PD-1/PD-L1 inhibition).

Certain pan-cancer clusters were suggestive of potentially novel relationships. For example, cluster 4 (defined by *CDKN2A* deletion) patients exhibited a pan-cancer reduction of PFS on cytotoxic chemotherapy (platinum-based training p-value = 4.78×10^-8, validation p-value = 1.92×10^-8; non-platinum-based training p-value = 7.22×10^-9, validation p-value = 2.76×10^-8). This trend was evident in non-small cell lung cancer, pancreatic cancer, and CUP samples (Figure 5). Of note, we examined CUP samples in this cluster with a high-confidence primary cancer type prediction (prediction probability >=0.5; Methods) using a previously published independently developed classifier specifically trained on multi-institutional mutational data from targeted sequencing assays to predict the most likely primary cancer type of tumor samples (OncoNPC)^55^. We observed that 50/90 (55.6%) samples were predicted to be either non-small lung cancer or pancreatic cancer. This again implies that the pan-cancer model may enhance power to detect associations across diverse clinical scenarios that may not be possible to assess with single-disease analyses. Whether there is a causal association between presence of a *CDKN2A* deletion and reduced response to cytotoxic therapy, or the observed association is explained by overall reduced prognosis for patients carrying this aberration^56^ is unknown. Nevertheless, there was limited evidence of reduced PFS on other therapy types associated with this cluster.

Another example is cluster 17 (defined by a combination of features, most strongly *APC* mutations, along with *FBXW7* missense mutations among others; Supplementary Table 9), which was associated with reduced PFS on PD-1/PD-L1 inhibition (training p-value = 6.02×10^-4; Figure 5). Prior work has shown an association between *APC* mutations and poor response to immunotherapy in colorectal cancer^57^, however, the trend we observed applied to samples from lung, endometrial, and esophagogastric cancers as well, and was only evaluable in the pan-cancer setting, suggesting a potential pan-cancer trend (Figure 5). Prior research has also established a mechanism of resistance to anti-PD1 therapy through inactivation of *FBXW7*^58^, which is mutated in 15.4% (2/13) of training set cases receiving PD-1/PD-L1 inhibitors in this cluster compared to 4.78% (70/1464) of cases receiving the therapy outside the cluster.

To globally assess the validity of the pan-cancer model, we again employed the Cauchy combination test^52^ which yielded a combined p-value of 5.5×10^-11 for the training set and 1.32×10^-6 for the validation set. Permutation testing of cluster assignments revealed a low probability of obtaining a Cauchy combined p-value at least as significant as observed in the model (p-value = 1.13×10^-4 based on 10,000 permutations). Together, these results suggest that the genomic based deep learning framework identified a clinically meaningful structure in the data.

### Using Pan-Cancer Patient Similarity Model in Precision Oncology Settings

We next assessed patient-specific neighborhoods for our training cohort (n=4,943), computing the percentage of samples with useful neighborhoods (AP at k=10 neighbors >= 0.2) as previously described (Methods; Supplementary Figure 1). In this cohort, 67.6%, 26.9%, and 12.9% of cases had significantly more useful neighborhoods than expected by chance when considering minimal, intermediate, or complete therapeutic overlap for neighbor relevance respectively (permutation p-values = 0.0002, 0.0002, 0.0024; Methods; Figure 6a). Identifying neighbors from the training set for the validation set cases (n = 3,195) demonstrated generalizability to unseen cases as 68.8%, 25.9%, and 12.1% of cases had useful neighborhoods with minimal, intermediate, or complete therapeutic overlap, respectively.

**Figure 6:**
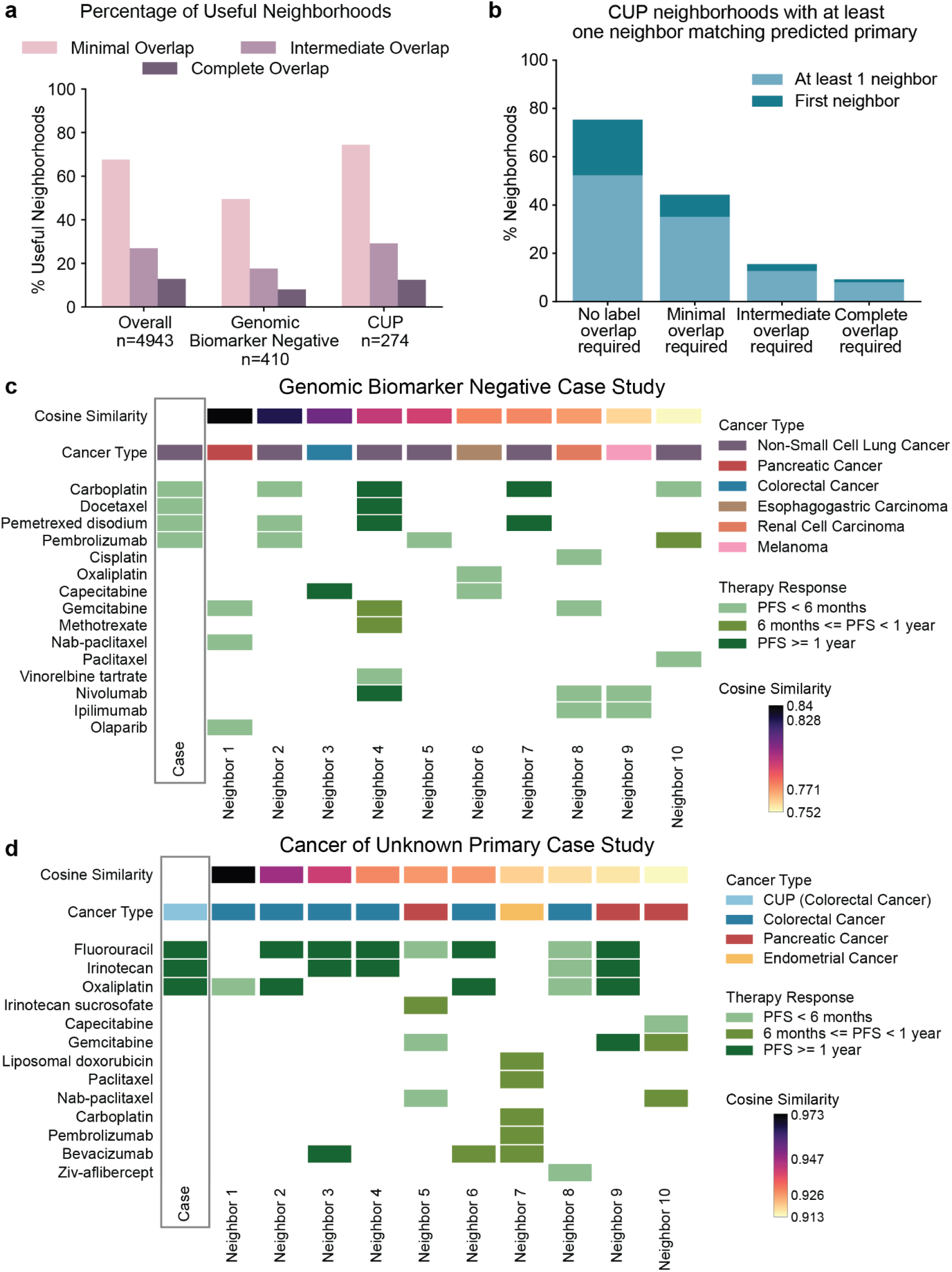
Pan-cancer Neighborhoods. a: Percentage of patients with useful neighborhoods in the pan-cancer cohort, as well the genomic biomarker-negative (Methods) and CUP sub-cohorts. Bar colors indicate the level of therapeutic overlap required to consider a neighbor as relevant (Methods). b: The percentage of CUP case neighborhoods where at least one of the 10 nearest neighbors matched the predicted cancer type of the CUP case (Methods). The darker color indicates the portion where the first neighbor matched the predicted primary. The first bar shows the percentage when that neighbor was not required to also share clinical labels with the case. The subsequent bars show the percentage when the neighbor sharing the predicted primary also shared clinical labels with the case under each level of overlap. c: Example of biomarker negative NSCLC sample with a useful neighborhood under complete therapeutic overlap (Methods), the neighbors’ clinical history may indicate the potential for poor response to chemotherapy and immunotherapy, prompting the clinician to consider different treatment options than what was ultimately pursued for the case patient. d: Example of a CUP sample with a useful neighborhood under complete overlap. The neighborhood may indicate to a clinician that the sample is likely a colorectal cancer sample, since it was the most common cancer type among the nearest neighbors. Indeed, the cancer type was also predicted as colorectal cancer by an independently developed model^55^ (Methods). The neighborhood may further indicate the potential for this patient to respond to chemotherapy.

Similar to the breast cancer-only model, we observed that clinically challenging cases could also have useful neighborhoods. Among 410 cases defined as genomic-biomarker negative in this cohort (Methods; Supplementary Table 7), 49.5%, 17.6%, and 8.04% of cases had useful neighborhoods under minimal, intermediate, and complete therapeutic overlap for neighbor relevance (permutation p-values not significant). Among CUP patients (n=274), 74.5%, 29.2%, and 12.4% of cases had neighborhoods with AP at k=10 neighbors >= 0.2 under minimal, intermediate, and complete therapeutic overlap, respectively (permutation p-value = 0.04 for minimal therapeutic overlap; not significant for intermediate and complete therapeutic overlap). These findings together demonstrate that a pan-cancer patient similarity framework may have utility for clinical decision making in some CUP cases and cases lacking actionable genomic biomarkers^55,59,60^ (Figure 6a).

We next investigated whether the sample neighborhoods in a pan-cancer environment could be diagnostically useful in identifying the most likely histologic source for CUP cases. We examined the subset of cases with a high confidence OncNPC^55^ prediction (prediction probability >=0.5) that matched one of the cancer types included in our pan-cancer model (Methods). In 45/174 cases (25.9%; permutation p-value = 0.0002), the most common cancer type among the neighbors matched the predicted primary cancer type of the case in question. 131 cases (75.3%, permutation p-value = 0.0002) had at least one neighbor from the same cancer type as the predicted primary, and 44.3%, 15.5%, and 9.2% of cases also shared therapeutic labels with minimal, intermediate, and complete therapeutic overlap respectively with at least one neighbor from the same cancer type as the predicted primary (permutation p-values = 0.0002, 0.0004, 0.0002; Methods; Figure 6b). These results underscore the potential utility of a generalized patient similarity framework applied to a pan-cancer patient population to guide clinical decision making specifically in CUP cases, as the sample neighborhoods may help in identifying the most likely primary diagnosis and indicate which therapies may be most relevant.

To further highlight the patient-specific applicability across cancer types, we visualized case studies that exemplify how the model may be used at the point of care (Figure 6c, d). One previously untreated patient with NSCLC was classified as genomic biomarker negative and had a useful patient neighborhood consisting of patients with reduced PFS to chemo-immunotherapies. This lack of response was ultimately the clinical trajectory this patient experienced as well. While the patient’s tumor did not have genomic biomarkers associated with FDA-approvals or clinical guidelines (Methods), it did harbor an *STK11* mutation that was shared with five of the neighbors and is consistent with emergent literature describing an association with reduced response to these therapy types^61,62^. A second patient with CUP had a useful neighborhood enriched for colorectal tumors, which would have further pointed to therapeutics employed in those settings; indeed, the patient ultimately received and responded to those therapies. In both example cases, information from patient neighborhoods, including histologic diagnoses and relative therapeutic outcomes to a variety of treatments, could have been provided to the clinician prior to decision-making to inform the index patient’s treatment considerations.

We also examined clinical scenarios in which tumors harbor multiple actionable genomic biomarkers that suggest distinct therapeutic strategies (Supplementary figure 7). The first involved a *ERBB2* amplified esophagogastric carcinoma case with high TMB (16.73 mutations/Mb). Current approvals would recommend trastuzumab based on the *ERBB2* amplification^47^ as well as pembrolizumab for high TMB^6^, yet only one of the nearest neighbors received pembrolizumab and exhibited a PFS less than six months, potentially guiding clinicians away from this option. The second case was an *EGFR* mutated NSCLC with high TMB (17.49 mutations/Mb). While the tumor characteristics might have presented competing therapeutic indications for *EGFR* and PD-1/PD-L1 inhibition, prior studies have shown reduced benefit to immunotherapy among *EGFR* mutant NSCLC cases^63–65^. Here too, the nearest neighbors treated with immunotherapy had short PFS, suggesting that alternative targeted strategies might be more appropriate, in line with current management of this disease subtype.

### Retraining the Model at Different Time points Reveals Stability and Tracks with Changes in Clinical Practice

Finally, to assess the stability and evolution of the framework results across time, and to simulate future integration and reevaluation of patient similarity modeling within a learning healthcare system, we tested how this approach would perform if trained at different time points using the data and clinical knowledge available at the time. We re-trained the breast cancer model 6 times, analyzing the structure it defined each year between 2017 and 2022. For each model, we only considered training set samples that were sequenced prior to the end of the corresponding year, and we used the contemporary version of the Molecular Oncology Almanac^1^ to create the input feature set (Methods).

The number of clusters identified at each iteration ranged between 10-12, with the 2022 model identifying the most clusters. We also found that the number of clusters we labeled as “genomic-biomarker specific” (Methods; Supplementary Table 10) fell between 7-9 clusters across models. Consistently in each time-frozen model, the number of samples assigned to clusters that were genomic-biomarker specific ranged between 59-71% and correlated with the number of “genomic-biomarker specific” clusters (Figure 7a, b). To measure the consistency of cluster assignments, we computed the adjusted mutual information (AMI) scores between cluster assignments from one year to the next for samples from the previous year. For example, the AMI score comparing the 2017 and 2018 models was based only on the samples that were already included in the 2017 model. We observed AMI scores in the range 0.568 to 0.667 (Supplementary Table 11), demonstrating a generally consistent mutual structure from one model to the next with some variation across the six year interval as new data was incorporated.

**Figure 7:**
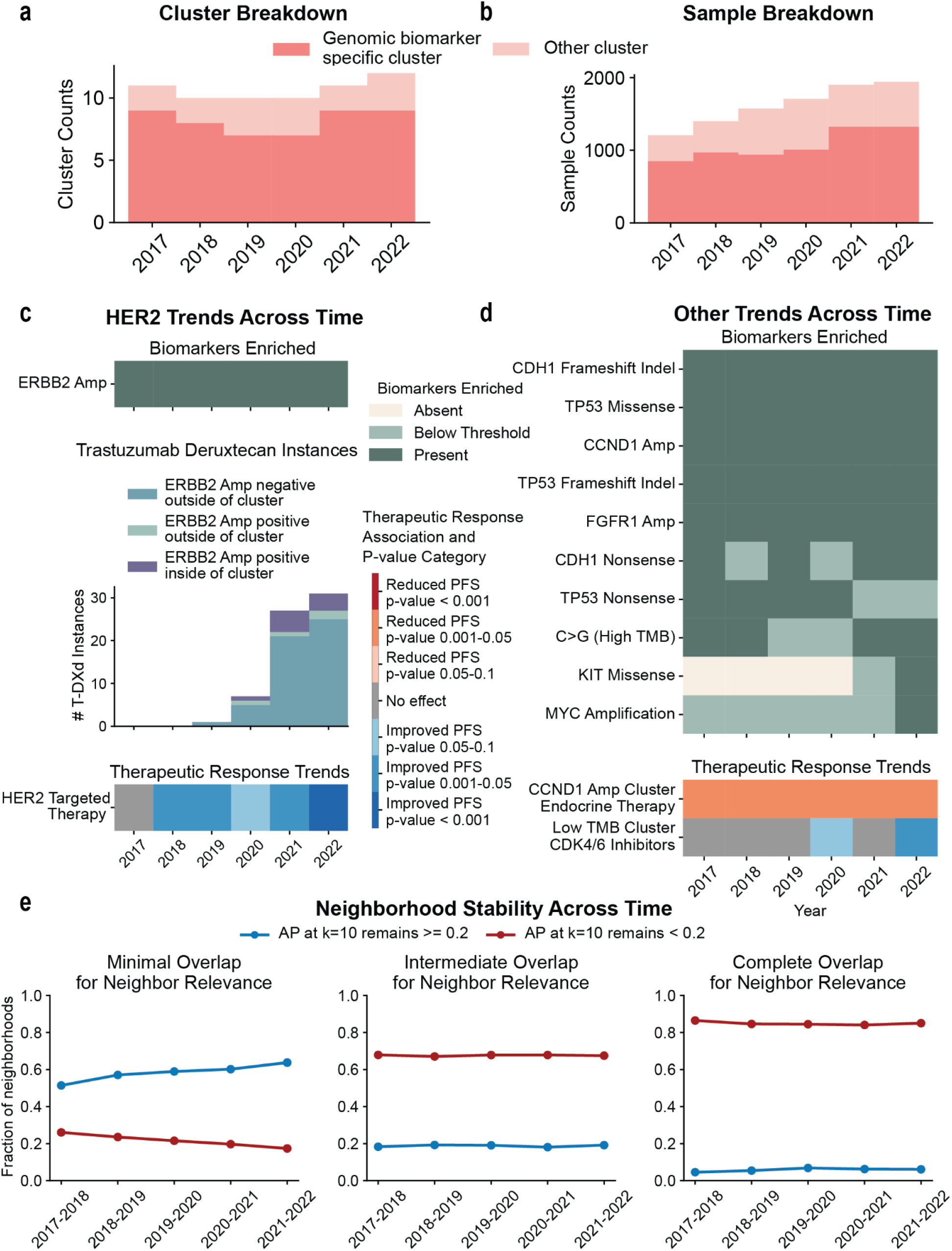
Retraining breast cancer model based on available data from 2017-2022. a: Observed cluster counts in each iteration, specifying “genomic biomarker specific” cluster counts (Methods). b: Sample counts in each iteration. Samples were categorized based on whether they belonged to a “genomic biomarker specific cluster”. c: Changes in HER2 specific trends across the model iterations. (Top) Presence of at least 1 cluster defined by *ERBB2* amplifications in each iteration. (Middle) Counts of patients receiving T-DXd whose associated sample was clustered inside vs outside of the *ERBB2* amplification cluster within each iteration. (Bottom) The p-value associated with improvement on HER2 targeted therapy for patients in the *ERBB2* amplification cluster within each model iteration. The darker blue indicates a lower p-value and gray indicates lack of trend with specified p-value thresholds. Note that the *ERBB2* amplification cluster in 2017 did show a non-significant trend of improved PFS on HER2 targeted therapy. d: Summary of other observed trends across the model iterations. (Top) Clusters defined by other genomic biomarkers (Methods). (Bottom) Key survival trends identified in the original breast cancer model across iterations. Blue depicts trends of improved PFS for patients receiving the indicated therapy type inside the cluster that’s defined by the indicated biomarker vs outside that cluster, and red shows trends of reduced PFS. The color shade indicates the range of the log-rank test p-value, with a darker shade indicating a lower p-value. Gray indicates lack of a trend with p-value < 0.1. Note that the low TMB cluster in 2021 did show a trend of improved PFS on CDK4/6 inhibitors, but it did not cross our p-value threshold. e: The stability of neighborhood classes from one model iteration to the next at each level of required overlap. Blue shows the fraction of neighborhoods that remain categorized as useful (AP at k=10 neighbors value >=0.2) after the update, red indicates the fraction of neighborhoods that remain categorized as not useful (AP at k=10 neighbors < 0.2).

We also computed how the trends identified by the models changed over time. For example, all models consistently identified an *ERBB2* amplification specific cluster, and the 2021 iteration identified two such clusters: one of which was specific to samples carrying a *TP53* missense co-mutation. We further observed that the signal of improved PFS on HER2-targeted therapy associated with the *ERBB2* amplification cluster was stably detected starting in 2018, and strengthened in 2022 (Figure 7c). Moreover, for the 2021 model, the response association was specific to the cluster defined only by *ERBB2* amplification, and not the *TP53* missense co-mutation cluster. The improvement of PFS over time was likely related to the incorporation of HER2-directed antibody drug conjugates (ADCs) into breast cancer clinical practice. In parallel with our observations, in 2019, trastuzumab emtansine (T-DM1) was FDA-approved as an adjuvant treatment of residual HER2+ breast cancer and T-DXd was approved for treatment of pre-treated, metastatic HER2+ breast cancer; both therapies significantly improved outcomes in HER2+ disease^66–69^. Furthermore, T-DXd was FDA-approved for earlier-line, metastatic HER2+ breast cancer in 2022, which may have driven the further improvement in PFS in *ERBB2*-amplified cases observed in our dataset^70^.

We subsequently evaluated the use of trastuzumab deruxtecan (T-DXd) inside and outside the *ERBB2* amplification enriched cluster across the model iterations. The first instance of use of the drug was in 2019, consistent with the timing of FDA approval in pre-treated, metastatic HER2+ disease^66,67,69^. The number of instances we observed in our data, which did not include therapies administered in clinical trials, steadily increased through our most recent available cases in 2022. Between 2019 and 2022, most of the instances of T-DXd usage were observed in *ERBB2* non-amplified cases outside the *ERBB2*-enriched cluster across all model iterations. Some of those patients may have been classified as HER2+ based on prior immunohistochemistry staining and receiving T-DXd as a second line HER2-directed therapy, while others may have been HER2-low and receiving T-DXd off-label in light of early reports of its efficacy in HER2-low disease in DESTINY-Breast04, which enrolled in 2018-2021^71,72^ (Figure 7c). Overall, these results showed that longitudinal analysis of the updating model reflected real-time trends in uptake and clinical benefit of novel therapies, despite model updates only incorporating genomic data from new samples.

Next we evaluated longitudinal trends captured by other genomic biomarker defined clusters (Figure 7d; Methods). Over the six-year period, most models consistently identified clusters associated with the same genomic biomarkers, although in certain cases that biomarker did not meet the pre-defined thresholds for linkage (Figure 7d; Supplementary table 10; Methods). For instance, *CDH1* nonsense mutations defined a cluster in 2018 but with an average value of 0.196, just below our inclusion threshold of 0.2 for this analysis, but above the threshold used in the breast cancer model. It was split between two clusters in 2020 and each remained below our threshold for linkage.

Regarding therapeutic trends identified by the models beyond the *ERBB2* amplification setting, trends with clinical evidence were also consistently identified at each model iteration for: (i) reduced PFS on endocrine therapy for samples in *CCND1*-defined clusters; and (ii) a low mutational burden cluster that was present in all models and for which we observed an association with improved PFS on CDK4/6 inhibitors, consistent with the previously unknown trend we described for the breast cancer model (Figure 7d). In the time-limited models, we began to see evidence of the low TMB cluster therapeutic trend in 2020, and this finding became statistically significant in 2022 (noting that we did observe the association in 2021, but it was below our pre-defined p-value threshold).

Lastly, anticipating scenarios where such models would be integrated into healthcare systems for real-time updates and applications, we assessed consistency in patient neighborhood utility across model runs. The percentage of cases whose neighborhoods changed from being useful (AP at k=10 neighbors >= 0.2) to not useful from one model iteration to the next ranged from 8.2-11.1% with minimal therapeutic overlap, 5.4-8% with intermediate therapeutic overlap, and 3.6-5.2% with complete therapeutic overlap. Similarly, the percentage of cases whose neighborhoods became useful from one model iteration to the next was 8.7-11.8%, 5.1-8.6%, and 3.6-5.3% under minimal, intermediate, and complete therapeutic overlap, respectively (Figure 7e; Supplementary Table 12). Thus, individual patients’ neighborhood utility for clinical decision support remained largely consistent across model updates, with some variations at the individual level. This suggests global stability of the framework, while allowing subtle locally adaptive changes within a real-time updating hospital setting as new data becomes available.

## Discussion

We developed a novel framework for evaluating patient tumor similarity and utilizing it as a generalized clinical decision-support tool that conveys complex information in a summarized and clinically useful format. Rather than serving as a stand-alone predictive model for a specific task, the framework identifies patient neighborhoods whose molecular profiles and clinical trajectories may be most relevant for the target patient. As studies on AI integration into clinical workflows underscore the risk of over-reliance on erroneous model outputs^73,74^, using AI to highlight relevant information, rather than generate predictions, may facilitate adoption while preserving clinician agency. Using our framework, we recovered expected clinically aligned patient groups, therapeutic similarity trends that mirror observations previously reported in literature, as well as others that have not been previously reported and may warrant further investigation. We showed that patient neighborhoods as assessed at the time of sequencing can inform the target patient’s therapeutic trajectory more often than expected by chance in both single disease and pan-cancer settings, supporting the potential use for clinical decision making aid at the point-of-care. The potential clinical utility of neighborhoods extended to patients with no known actionable genomic biomarkers or with a CUP diagnosis, providing guidance for complex diagnostic scenarios and for patient groups who currently face limited therapeutic outcomes. We demonstrated that the neighborhoods of CUP patients were significantly enriched for samples that aligned with the predicted primary based on an independently developed model^55^, underscoring the flexibility of our approach (which was not specifically and exclusively intended for CUP inference) to provide context-specific decision support at the clinician’s discretion. Broadly, our approach offers a methodological precision oncology framework, where heterogeneous genomic and clinical data are distilled into clear, context-specific insights that augment, rather than replace, clinical expertise.

However, we acknowledge several limitations. First, while we use a held out validation set to demonstrate the generalizability of the learned embeddings and cluster assignments to unseen samples, the data was all obtained from a single tertiary cancer center, and the trained model may not be directly generalizable to data from other institutions. In agreement with recent discussions in the field^75–77^, we believe that given differences in sequencing assays and clinical data collection protocols in different institutions, the framework would need to be independently trained for deployment elsewhere, and we provide the full details of our methodology for this purpose. We expect this limitation of the field to be alleviated as large, consortia efforts make progress towards harmonizing clinicogenomic data from multiple institutions^78^.

Second, our sample embeddings relied on genomic data generated using a targeted sequencing assay, enabling us to optimize genomic embeddings primarily based on exonic mutations, focal copy number alterations, and fusions in the targeted genes, without being able to measure genome wide features, broad copy number alterations, or non-genomic biomarkers. We envision future extensions of this work to utilize whole-exome or whole-genome sequencing data, and to incorporate orthogonal data modalities such as histopathology images, immunohistochemistry staining, laboratory results, and clinical history. Such a multimodal co-embedding would enable identification of similar prior patients based on a more comprehensive feature set, and may then be further fine-tuned for specific downstream predictive tasks.

Furthermore, this study utilized real-world data, and thus it suffers from heterogeneity among samples and missing data. For instance, our clinical data was limited to medications received at the cancer center outside of clinical trials, potentially hindering our downstream analysis of clinical similarity due to incomplete treatment histories for some patients. In addition, the progression times used in our analyses were derived from downstream processing of natural language processing (NLP)-extracted annotations from clinical and imaging reports^45^ (Methods) and are therefore subject to potential errors. Our data also included patients at different stages of disease progression and treatment, and is subject to biases that commonly affect precision oncology studies: that the time of genomic sequencing correlates with risk and may itself be an indication of a progression event^79,80^.

Finally, we emphasize that the observed associations do not establish causal relationships and that we cannot rule out confounders. While we assessed the significance of our results above random patient groups and controlled for various confounding variables that may have driven our results, there may be additional latent variables unknown to us. We assessed the significance of neighborhood utility in the genomic embeddings space relative to what was expected by chance using permutation analyses in which we accounted for the structure contribution driven by samples from the same patient or the same cancer type. Still, additional confounding structure may have contributed to the observed signal, potentially inflating the observed model utility.

Our tumor similarity model offers a framework for more individualized treatment guidance by considering the mutational profile of each patient’s tumor and drawing on prior patient experiences. This framework represents an information-retrieval-based AI approach that aligns with clinician-centered workflows and lends itself to generalized use across varied clinical settings and applications. By utilizing real-world data, our approach can further advance precision oncology by deriving holistic biomarkers in real-life clinical settings that may differ from randomized clinical trials^81–84^. We demonstrated the stability of our model when retrained at different time points, suggesting its potential as a real-time self-learning precision oncology framework that can be embedded into a healthcare system. Agentic implementation within the electronic health record may offer a comprehensive way to continuously digest these large quantities of complex data to enhance next generation patient similarity approaches for precision oncology.

## Author contributions

M.S., B.R., and E.M.V.A designed the study. M.S. processed the data, developed and implemented the model, and performed analyses. B.R assisted with data processing and model development and analyses. S.J., E.P., and T.O assisted with analyses. E.P. and T.O assisted with clinical interpretation of results. J.P. assisted with writing and editing the manuscript. K.L.K provided the data and assisted with data processing and result interpretation. H.A.E assisted with the model evaluation. S.R.S assisted with statistical analyses, and provided guidance and feedback. M.S. and E.M.V.A wrote the manuscript. All authors read, edited, and approved the final manuscript.

## Acknowledgements

This work was funded by the National Institutes of Health (NIH) National Cancer Institute (NCI) grant R01CA278980 to E.M.V.A, Department of Defense (DOD) grants W81XWH-21-PCRP-DSA and HT94252410415 to E.M.V.A., Mark Foundation Emerging Leader award to E.M.V.A, and NIH National Institute of General Medical Sciences (NIGMS) grant 5R35GM127131-09 to S.R.S. M.S. was partially funded by the Eric and Wendy Schmidt Center (EWSC) at the Broad Institute of MIT and Harvard. J.P. was funded by NCI grant R50CA265182. H.A.E was funded by the DOD CDMRP grant HT9425-23-1-0023. Schematics in this paper were created in Biorender.

## Data Availability

The underlying EHR data used for these analyses constitute protected health information for DFCI patients and therefore cannot be made publicly available. Researchers with DFCI appointments and Institutional Review Board (IRB) approval can access the data on request. For external researchers, access would require collaboration with the authors and eligibility for a DFCI appointment per DFCI policies. Deidentified genomic data are available for DFCI patients through AACR’s Project GENIE. Deidentified clinical data corresponding to PRISSMM annotations, pathologic information, imaging information, and medical oncologist evaluation for some DFCI patients is publicly available on cBioPortal and Synapse through the AACR Project GENIE Biopharmaceutical Consortium.

## Declaration of generative AI and AI-assisted technologies in the writing process

During the preparation of this work the authors used ChatGPT for sentence refinement. After using this tool, the authors reviewed and edited the content as needed, and take full responsibility for the content of the published article. All substantive content was written, reviewed, and approved by the authors.

## Methods

### Data Processing

#### Dataset

The study utilized data for patients treated at Dana-Farber Cancer Institute whose tumors were profiled using the OncoPanel hybrid-capture sequencing assay^5^ between 2013 and 2024. We only considered tumor samples with a reported tumor purity of at least 20% for model training.

Clinical data for these patients were extracted from electronic health records (EHR). Downstream clinical analysis was limited to patients for whom we have information on last known alive date or date of death, at least one imaging report and medical oncology report, and at least one cancer medication entry given within two years after the tumor sample sequencing date. To identify cancer-targeting medications, we identified instances of medications given under a standard chemotherapy plan according to our data, removing supportive medications and medications given under a clinical trial. To account for incomplete or erroneous treatment plan annotations, we further curated the medication list by referencing the Molecular Oncology Almanac^1^ and consulting medical oncologists to ensure medications included were truly limited to cancer-targeting medications and to annotate their corresponding therapy strategy (Supplementary Table 13).

Patients were split into training (60%) and validation (40%) subsets, and all model development was limited to samples from patients in the training set. Patient splits were stratified to maintain similar distributions of the OncoPanel version used and the biopsy type (primary or local recurrence vs metastatic recurrence) of the included samples across the subsets. The breast cancer model included all samples satisfying the above criteria from patients who have had a breast cancer primary diagnosis and consisted of 2,231 samples from 2,097 patients in the training cohort and 1,480 samples from 1,399 patients in the validation cohort. Therapeutic labels were available for 993 and 662 patients in the training and validation sets respectively. The pan-cancer training cohort consisted of 17,615 sequenced samples from 16,522 patients with therapeutic labels available for 5,705 patients. The validation cohort consisted of 11,804 samples from 11,055 patients with therapeutic labels available for 3,723 patients. The pan-cancer model broadly consisted of solid tumors, specific cancer types included were: breast cancer (BRCA), ovarian cancer (OV), endometrial cancer (UCEC), prostate cancer (PR), thyroid cancer (THYR), non-small cell lung cancer (NSCLC), small cell lung cancer (SCLC), head and neck carcinoma (HNC), esophagogastric Carcinoma (EGC), bladder cancer (BLCA), pancreatic cancer (PAAD), hepatocellular carcinoma (HPC), biliary cancer (BIL), gastrointestinal stromal tumor (SGT), renal cell carcinoma (RCC), colorectal cancer (COADREAD), melanoma (MEL), other skin cancers (SK), and cancer of unknown primary (CUP) (Supplementary Table 2).

#### Genomic Data Processing

Tumor somatic profiles were sequenced using the OncoPanel hybrid-capture sequencing assay^5^. The genes captured with the OncoPanel assay have been updated based on emerging clinical or biological knowledge, resulting in three different versions of the assay used, with additional genes included in each version during the time period considered. To maximize the number of samples in our training set, we focused on the shared genes across all three versions, which is 241 genes (Supplementary Table 14). The genomic data included single nucleotide variants (SNVs) and insertions/deletions (indels), focal copy number amplifications and deletions (CNVs), and structural variants (SVs).

To extract feature vectors from the mutation data for model training, we relied on annotations of molecular alteration clinical actionability from the Molecular Oncology Almanac (MOAlmanac) database version 2022-03-03^1^. SNVs and indels were first filtered to remove possible germline variants by excluding all variants that appeared in the Exome Aggregation Consortium (ExAC) with a minimum allele count of 10, with the exception of loci where known pathogenic somatic variants may occur^85,86^. We designed the feature set extracted from the remaining variants with the goal of capturing global properties of the tumor genomic profiles, while ensuring potentially actionable variants are taken into account. As such, we included a set of indicator features for the status of specific alterations related to FDA-approvals or clinical guidelines, according to the MOAlmanac database as well as mutation counts in each of the 241 genes, partitioned by variant type. In addition, we included a feature for overall coding mutation burden, defined as mutations with a variant classification of ‘frameshift indel’, ‘inframe indel’, ‘missense’, ‘nonsense’, ‘nonstop’, ‘splice site’, and ‘translation start site’; in addition to single base substitution (SBS) counts. For copy number variations (CNV) and structural variations (SV), we included features for genes associated with any level of evidence in the MOAlmanac database. CNV features were encoded as two features per gene indicating the presence or absence of either an amplification or a deletion of that gene, requiring a log fold-change of 2 to consider an alteration as present. SV features were encoded as the presence or absence of a fusion involving at least one of the included genes, with fusions involving two included genes appearing twice in the feature vector. For each model trained, we ignored features that were zero cohort-wide in the training set. The final feature vector size was 1071 for the breast cancer model and 1452 for the pan-cancer model (Supplementary Table 15).

Prior to training or applying the model, the feature matrix was normalized using min-max normalization, with normalization parameters set based only on the training set to avoid information leakage.

#### Clinical Data Processing

The EHR data we utilized for our patient cohort included natural language processing (NLP)-derived progression labels from medical oncology notes and radiology reports from an independently developed previously published model^45^. In this work, a subset of imaging reports and medical oncologist notes were manually annotated for several outcomes, including disease progression and response to therapy based on the Pathology, Radiology/Imaging, Signs/Symptoms, Medical oncologist assessment, and bioMarkers (PRISSMM) framework^87^, and a transformer-based model was trained to predict those annotations. We also extracted the patient’s last known alive date, and date of death if applicable, as well as information on medications received.

We represented patient clinical trajectories as progression-free survival (PFS) on oncology-related medications received within two years after sample sequencing, grouped into therapeutic strategies. As discussed in the data inclusion section above, we only focused on medications that were administered within a standard oncology treatment plan, excluding medications given within clinical trials as well as supportive medications that aimed to mitigate side effects of the cancer treatment. The raw medication data listed individual intravenous (IV) medication administration sessions and prescriptions for oral medications for each patient. To process the remaining data into single-agent regimens received by the patient, we combined IV medication sessions or prescriptions of the same medication that occur within 120 days of one another into single-agent regimens that started on the date of the first instance of the medication. We grouped medications into therapy strategies that included targeted therapies (e.g. EGFR inhibitors, BRAF inhibitors, etc), hormone therapies (i.e. antiandrogens, ER signaling inhibitors, and aromatase inhibitors), immunotherapies (i.e. PD-1/PD-L1 inhibitors and CTLA-4 inhibitors), and chemotherapies (i.e. cytotoxic therapies separated into platinum and non-platinum based).

To compute each patient’s progression-free survival (PFS) on each medication, we considered medical oncology notes and radiology reports occurring at least two weeks after the processed medication start date. We recorded a progression event to have occurred if it was indicated by a medical oncology and radiology report within 30 days of one another according to the NLP model predictions. If a progression event was found with these criteria, we set the event date as the later date of the two reports. If no progression event was observed according to these criteria, then we considered the patient’s date of death if applicable or their last known alive date in computing the PFS.

### Model Training

#### Creating Sample Genomic Embeddings

To create sample genomic embeddings we trained a self-supervised autoencoder model (Supplementary Table 16) with encoder and decoder networks that consist of 2 hidden layers each. Training was performed for 2000 epochs on a batch size of 128, and the embedding layer consisted of 50 nodes for the breast cancer model and 128 nodes for the pan-cancer model. Hyperparameters tuned included learning rate and activation functions, with the final models utilizing a learning rate of 0.0001; and leaky ReLU activation for the hidden layers, and ReLU activation for the reconstruction layer. Cross-validation was used to ensure consistency in convergence patterns and for hyperparameter selection. The loss function used was mean squared error (MSE). Once trained, the final model was used to create embeddings for the training and validation sets. The model was implemented using Pytorch version 1.12.1.

We additionally experimented with other models for developing a similarity measure in the genomics space: 1) K-means clustering and K-NN directly on un-embedded genomic features (see below); 2) PCA transformation to create a lower-dimensional representation of genomic features prior to clustering and neighborhood analysis; 3) training a variational autoencoder (VAE) to create the genomic embeddings. The VAE was trained using a similar architecture and hyperparameters as the autoencoder model. To compare these models we evaluated the cluster size distribution and the generalizability of cluster assignment to the validation set. We found the autoencoder model to be the most generalizable (Supplementary Table 1)

#### Identifying Clusters in the Genomic Embedding Space

K-means clustering was used to identify sample clusters in the genomic embedding space (or based on unembedded genomic features for model comparisons). The optimal number of clusters was selected within a pre-specified range (10-20 for the breast cancer model and 10-30 for the pan-cancer model) by optimizing Silhouette score. Clusters were identified based on all training samples for each cohort, regardless of clinical label availability, and applied to all validation samples. This was implemented using Scikit-learn version 1.0.2.

#### Identifying Nearest Neighbors Based on Genomic Embedding

To explore the relationship between the genomic similarity and the patient clinical trajectories, we computed the 10-nearest neighbors in the genomic embedding space using cosine-similarity as the distance metric. To simulate realistic clinical use of the model, we limited the search space for each target sample’s neighborhood to the training samples with available clinical data that were sequenced prior to the target sample. To evaluate the utility of the neighborhoods, we only considered clinical labels that were available at the time of the query sample’s sequencing as will be discussed in the evaluation section. To assess generalizability to unseen patients, we also computed the 10-nearest neighbors for each of the validation samples among the previously sequenced training samples with available clinical data, and evaluated the neighborhood utility using the same procedure (described below).

### Model evaluation

#### Evaluation of Prognostic Value of Genomic Clusters

To examine the clinical trends identified by our clusters, we used a Kaplan-Meier survival estimator to compare samples inside each cluster with samples outside the cluster across various therapy types using a log-rank test. The analysis included only cluster–therapy strategy pairs with at least 10 training-set samples both inside and outside the cluster that received the therapy. We considered only the first instance of a given therapy strategy following sample sequencing. For each model, we performed the analysis on the entire training cohort as well as on subcohorts. The subcohorts considered were: 1) cases where the therapy intent was not curative, adjuvant, or neo-adjuvant (referred to as non-curative intent therapies in the main text); 2) cases where the therapy analyzed was the first therapy line given according to our data; 3) cases where the patient had received at least 1 prior line of therapy; 4) only biopsy samples from primary tumors; 5) only biopsy samples from locally recurrent or metastatic tumors; and, in the case of the pan-cancer model, 6) only on cases from each specific cancer type. The goal of the subcohort analyses was to assess whether the observed difference on PFS was specific to those sub-cohorts, and the minimum sample counts per category required for these stratified analyses was 5 instead of 10. We also performed the analyses separately on the validation set with the same minimum sample requirements to assess generalizability of the observed associations. The analysis was implemented using the Lifelines Python package version 0.27.7.

To address the issue of multiple testing in this analysis and test the reliability of observed cluster-associated survival trends, we ran the Cauchy combination test^52^ on the log-rank test results for the training and validation sets separately. The Cauchy combination test assesses the presence of a significant signal in the data without assuming independence of individual tests, making it appropriate for our analysis. In the case of the breast cancer model, we also ran the test after removing the expected relationship of improved PFS on HER2 targeted therapy for samples in the *ERBB2* enriched cluster to assess the presence of a significant trend in the data beyond this well-validated association. Furthermore, to assess whether the structure identified by our model associated with a stronger clinical signal than expected by chance, cluster assignments were permuted 10,000 times, and the full survival analysis was repeated for each permutation to estimate the probability of obtaining an equal or lower Cauchy combined p-value by chance in the training set.

To interpret these trends, we trained L1-regularized logistic regression models to predict cluster membership based on raw genomic features, and examined the coefficients assigned to each input feature for each model. To identify “cluster-defining biomarkers” for each cluster we considered features assigned a high positive coefficient (>=2.5) for predicting membership of that cluster and that have an average value among the training samples in the cluster of at least 0.1. For visualizing these biomarkers, we used higher thresholds for the pancancer model (logistic regression coefficient of at least 5 and average value among training set samples of at least 0.2) for ease of visualization due to the higher number of clusters. The models were trained only on the training set samples and validated on the validation set to ensure generalizability of the latent space and cluster assignment interpretation. Evaluation was done by computing the area under the receiver operating curve (AUROC) and the area under the precision-recall curve (AUPRC). High TMB clusters were identified based on coding mutation burden related features used in our model (total coding mutation count and individual SBS counts). Training set samples in the high TMB cluster in the breast cancer model (cluster 6) had an average coding burden of 23.12 compared to an average of <= 7.16 in other clusters, and training set samples in the high TMB cluster in the pan-cancer model (cluster 12) had an average coding burden of 48.29 compared with <= 10.43 in other clusters. Low TMB clusters were defined by an average coding burden of ∼2, compared with an average of >=4 across all other clusters in the same model, and a negative coefficient for a TMB-related feature in the cluster-membership predictive model. The logistic regression models were implemented using Scikit-learn version 1.0.2.

#### Evaluating Neighborhood Utility In Aiding Clinical Decision Making

To evaluate the model’s utility as a clinical decision-making aid for individual patients at the point of care, we analyzed each sample’s neighborhood as defined by the 10-nearest neighbors in the embedding space among previously sequenced samples, requiring at least 100 previously sequenced samples to consider (n=795 for the breast cancer model and n=4943 for the pancancer model). We excluded patients with multiple sequenced samples from this analysis to avoid overestimating the aggregate cohort-level neighborhood performance metrics.

To evaluate each patient’s neighborhood we utilized average precision metrics used to evaluate information retrieval systems as described previously^1^. To compute average precision of the 10 nearest neighbors, we defined each sample’s labels based on therapy classes of medications received and PFS range on each therapy class (e.g. PFS <= 6 months on Aromatase Inhibitors) and used three levels of stringency to define which neighbors were “relevant” for informing the case’s clinical trajectory. The most permissive level (“minimal” therapeutic overlap) considered a neighbor to be relevant if they shared at least one label with the case, the second level (“intermediate” therapeutic overlap) considered a neighbor to be relevant if they shared all of the case’s labels even if the neighbor received additional therapies not received by the case, and the strictest level (“complete” therapeutic overlap) considered a neighbor to be relevant only if their labels were exactly the same as the case’s labels (Figure 1c, Supplementary Figure 1). This allowed us to compute three values for each case’s neighborhood utility. We then computed the average precision values across the 10 nearest neighbors (AP at k=10 neighbors) of each case in the cohort and determined the percentage of cases whose neighborhood’s AP at k=10 neighbors was at least 0.2. Neighborhoods that had an AP at k=10 neighbors of at least 0.2 were referred to as “useful” in this study. This threshold was defined in accordance with the average precision value of the best performing model for tumor to cell line matching reported by Reardon et al^1^. We note that this threshold was set prior to the analysis and we did not consider other thresholds. To assess the generalizability of the results to unseen patients, we computed and evaluated neighborhoods from among the previously sequenced training set samples for each patient in the validation set, again limiting to cases for which there were at least 100 previously sequenced samples in the training set (n=522 for the breast cancer model and n=3195 for the pancancer model)

To evaluate whether the cohort-wide neighborhood utility obtained by the model exceeded what was expected by chance, we implemented permutation tests to estimate the null neighborhood utility. We permuted the sample names associated with the embeddings 5000 times within each cancer type, maintaining the original sample name association with the therapeutic labels, and recomputed neighborhoods and neighborhood metrics. These permutations effectively led to randomly selected neighbors for each case, while controlling for cancer type to prevent a false positive result as a consequence of breaking the cancer type structure in the neighborhoods. We then computed the p-value associated with the model’s neighborhood utility as the percentage of permutations with at least as many useful neighborhoods as the model for each level of therapeutic overlap. Specifically, 𝑝 𝑣𝑎𝑙𝑢𝑒 = (𝑥 + 1)/(𝑝𝑒𝑟𝑚𝑢𝑡𝑎𝑡𝑖𝑜𝑛 𝑐𝑜𝑢𝑛𝑡 + 1), where 𝑥 was the number of permutations with neighborhood utility at least as good as the model.

We also evaluated the utility of neighborhoods of genomic biomarker negative cases specifically (n=263 and 410 for the breast cancer and the pancancer cohorts, respectively). To identify genomic biomarker negative cases, we considered the 2024-12-31 version of the MOAlmanac database, utilizing entries with a predictive implication of either “FDA-Approved” or “Guideline”^1^. Biomarker negative cases were defined as all samples lacking any SNV, indel, CNV, or SV that’s indicated with those predictive implications in the database. To account for entries that did not specify a specific alteration or variant type, biomarker negative cases were defined as samples lacking any SNV or indel in those genes. For the breast cancer cohort, we only considered MOAlmanac entries that are associated with breast cancer, but for the pancancer cohort we considered all entries regardless of the associated cancer type. Samples with an OncoPanel reported MMR deficient status (dMMR) or with OncoPanel reported tumor-mutational burden (TMB) of at least 10 mutations per megabase were defined as biomarker-positive for both cohorts, noting that those labels were not available for all samples.

To evaluate whether cancer of unknown primary (CUP) sample neighborhoods could aid in diagnosing the primary cancer type, we utilized predictions from a previously published independently developed model trained on somatic data from targeted sequencing assays to predict the most likely primary cancer type of tumor samples (OncNPC) as a ground truth^55^.

Focusing specifically on samples for which the OncNPC model predicted a primary cancer type that was included in our pan-cancer cohort with a probability of at least 0.5, we evaluated how often at least one of the 10 nearest neighbors matched the case’s predicted primary cancer type and how often the neighborhood’s majority cancer type matched the prediction. To further assess how often the therapeutic trajectory of neighbors with a matching cancer type could have informed the CUP case’s therapeutic trajectory, we computed how often at least one neighbor of a matching cancer type to the predicted primary would be considered relevant under each level of therapeutic overlap described previously. To evaluate the significance of these results, we also computed these metrics in each of the permutations described earlier.

To visualize the neighborhoods and perform case studies, we utilized the Python CoMut package^88^. In these visualizations, we included individual medications instead of aggregating them into therapy strategies, and we only included the first instance of each medication for each sample for clarity of visualization. The therapeutic information included for each patient in the neighborhood was censored by the query case’s sequencing date to simulate the information that would be available to clinicians at the point of care.

### Training and Evaluating Time-Limited Versions of the Model

Six time-limited breast cancer models representing the years 2017-2022 were trained using the same model architecture and hyperparameters as the original models, but with data limited to what was available by the last day of the corresponding model year. For example, the 2017 model was trained only on samples from the original training set that were sequenced prior to 2017-12-31, and with feature extraction done using the same procedure as for the original model but based on the 2017-12-31 version of MOAlmanac^1^.

To define “genomic biomarker specific” clusters and identify the genomic features explaining each cluster, we used the same thresholds described above for visualizing cluster-defining biomarkers in the pan-cancer model. Specifically, we identified features with logistic regression coefficients ≥5 for predicting each cluster’s membership and an average value ≥0.2 among training samples in the corresponding cluster. We utilized the higher thresholds in this case for ease of visualization. Any cluster for which we identified at least one feature satisfying these thresholds was considered a genomic-biomarker specific cluster. The low TMB cluster for each model was again defined by an average coding burden of ∼2, compared with a minimum average of ∼4 across all other clusters in that model, and a negative coefficient for a TMB-related feature in the cluster-membership predictive model.

The survival analysis for each model followed the same procedure described above. To identify specific therapeutic associations in each model, we tested whether the cluster defined by the genomic biomarker for each trend exhibited the same PFS difference observed in the breast cancer model (as assessed by a hazards ratio in the expected direction and a log-rank test p-value within one of the pre-defined ranges: <0.001, 0.001-0.05, or 0.05-0.1). In cases where more than one cluster in a specific model were enriched for the associated biomarker (according to the criteria described above), we checked if any of them showed the expected trend.

To assess consistency in cluster assignments or neighborhood utility across the time-limited models, we compared each model to the next based on the training samples from the previous year. For example, in comparing the 2017 model to the 2018 model results, we considered the cluster assignments and neighborhood AP at k=10 neighbors of the training set samples that were present in 2017; and for the 2018 to 2019 model comparisons, we considered all training set samples that were present in 2018; and so on. To evaluate cluster assignment consistency, we computed the adjusted mutual information score between assignments from one year to the next using the Scikit-learn library version 1.0.1. To evaluate consistency in neighborhood utility, we computed the percentage of samples whose neighborhood utility label (“useful” if average precision at k=10 neighbors was at least 0.2), remained consistent from one year to the next.

