## Supplementary Figures for "Learning Patient Similarity from Genomics for Precision Oncology"

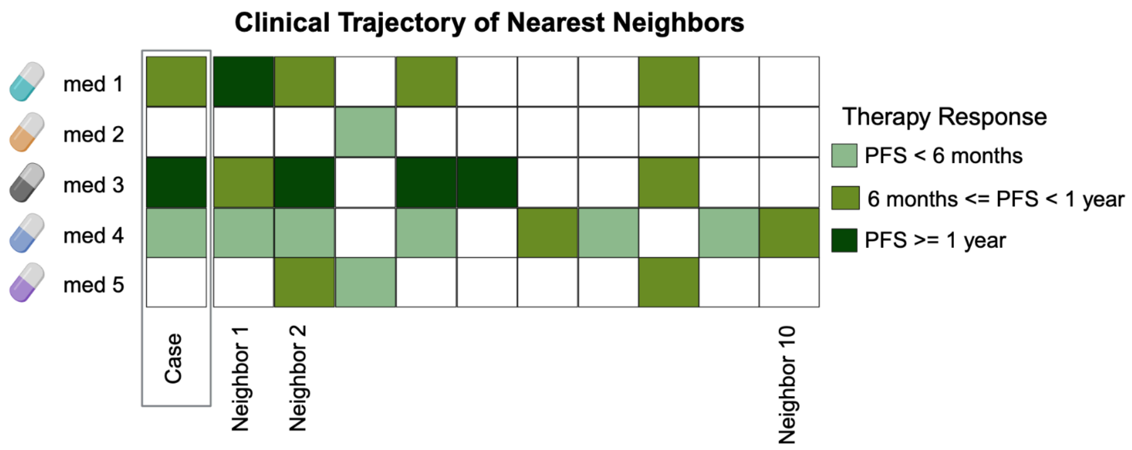


**Minimal Overlap**

| | | | | | | | | | | |
|---|---|---|---|---|---|---|---|---|---|---|
| Relevance@k | 1 | 1 | 0 | 1 | 1 | 0 | 1 | 1 | 1 | 0 |
| Precision@k | 1/1 | 2/2 | 2/3 | 3/4 | 4/5 | 4/6 | 5/7 | 6/8 | 7/9 | 7/10 |

Average Precision at 10 Neighbors = (1/count relavent) x Σ(relavence@k x precision@k)

= (1/7)(1+1+3/4+4/5+5/7+6/8+7/9) = 0.827

**Intermediate Overlap**

| | | | | | | | | | | |
|---|---|---|---|---|---|---|---|---|---|---|
| Relevance@k | 0 | 1 | 0 | 1 | 0 | 0 | 0 | 0 | 0 | 0 |
| Precision@k | 0/1 | 1/2 | 1/3 | 2/4 | 2/5 | 2/6 | 2/7 | 2/8 | 2/9 | 2/10 |

Average Precision at 10 Neighbors = (1/2)*(1/2 + 2/4) = 0.5

**Complete Overlap**

| | | | | | | | | | | |
|---|---|---|---|---|---|---|---|---|---|---|
| Relevance@k | 0 | 0 | 0 | 1 | 0 | 0 | 0 | 0 | 0 | 0 |
| Precision@k | 0/1 | 0/2 | 0/3 | 1/4 | 1/5 | 1/6 | 1/7 | 1/8 | 1/9 | 1/10 |

Average Precision at 10 Neighbors = (1/1)*(1/4) = 0.25

Supplementary Figure 1: Schematic of neighborhood average precision (AP at k=10 neighbors) calculation. The nearest neighbors to each patient are evaluated for their relevance, defined by whether they have a PFS in the same range on the same administered therapy types as the case patient. This is done at three stringency levels: “minimal” overlap considers a neighbor to be relevant if they share at least 1 label (i.e. have a similar PFS on at least 1 therapy type); “intermediate” overlap considers a neighbor as relevant if they share all of the case’s labels but may have additional therapies not represented in the case trajectory; “complete” overlap considers a neighbor relevant only if they have an identical trajectory to the case. This is summarized into an average precision score for each patient’s neighborhood at each level of stringency.

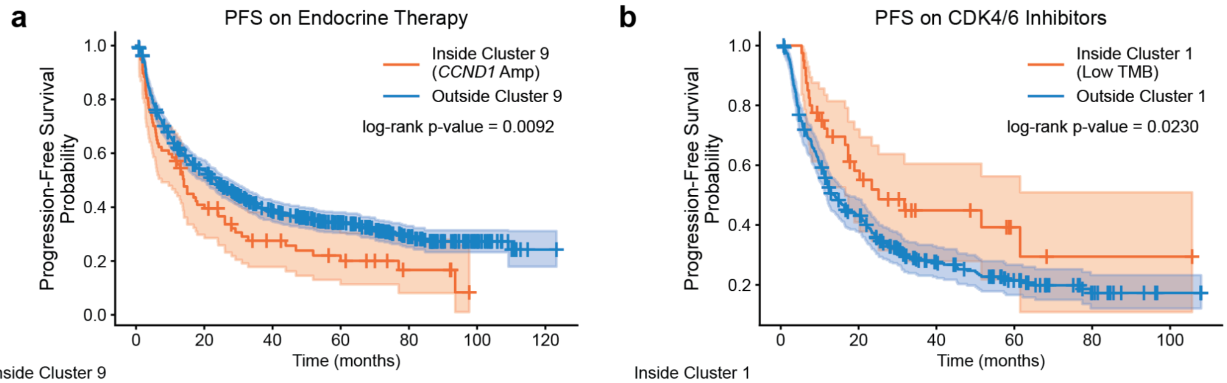


| Inside Cluster 9 (*CCND1* Amp) | | | | | | | |
|---|---|---|---|---|---|---|---|
| At risk | 77 | 30 | 16 | 10 | 4 | 0 | 0 |
| Censored | 0 | 2 | 7 | 9 | 14 | 17 | 17 |
| Events | 0 | 45 | 54 | 58 | 59 | 60 | 60 |
| **Outside Cluster 9** | | | | | | | |
| At risk | 667 | 343 | 209 | 146 | 74 | 25 | 1 |
| Censored | 0 | 19 | 61 | 103 | 157 | 202 | 225 |
| Events | 0 | 305 | 397 | 418 | 436 | 440 | 441 |

| Inside Cluster 1 (Low TMB) | | | | | | |
|---|---|---|---|---|---|---|
| At risk | 40 | 19 | 9 | 4 | 1 | 1 |
| Censored | 0 | 5 | 11 | 15 | 17 | 17 |
| Events | 0 | 16 | 20 | 21 | 22 | 22 |
| **Outside Cluster 1** | | | | | | |
| At risk | 322 | 131 | 59 | 30 | 11 | 2 |
| Censored | 0 | 12 | 40 | 57 | 72 | 81 |
| Events | 0 | 179 | 223 | 235 | 239 | 239 |

Supplementary Figure 2: Survival trends in breast cancer model. a: Reduced PFS on Endocrine therapy for samples in the *CCND1* amplification defined cluster (cluster 9). b: Improved PFS on CDK4/6 inhibitors for samples in the low TMB cluster (cluster 1).

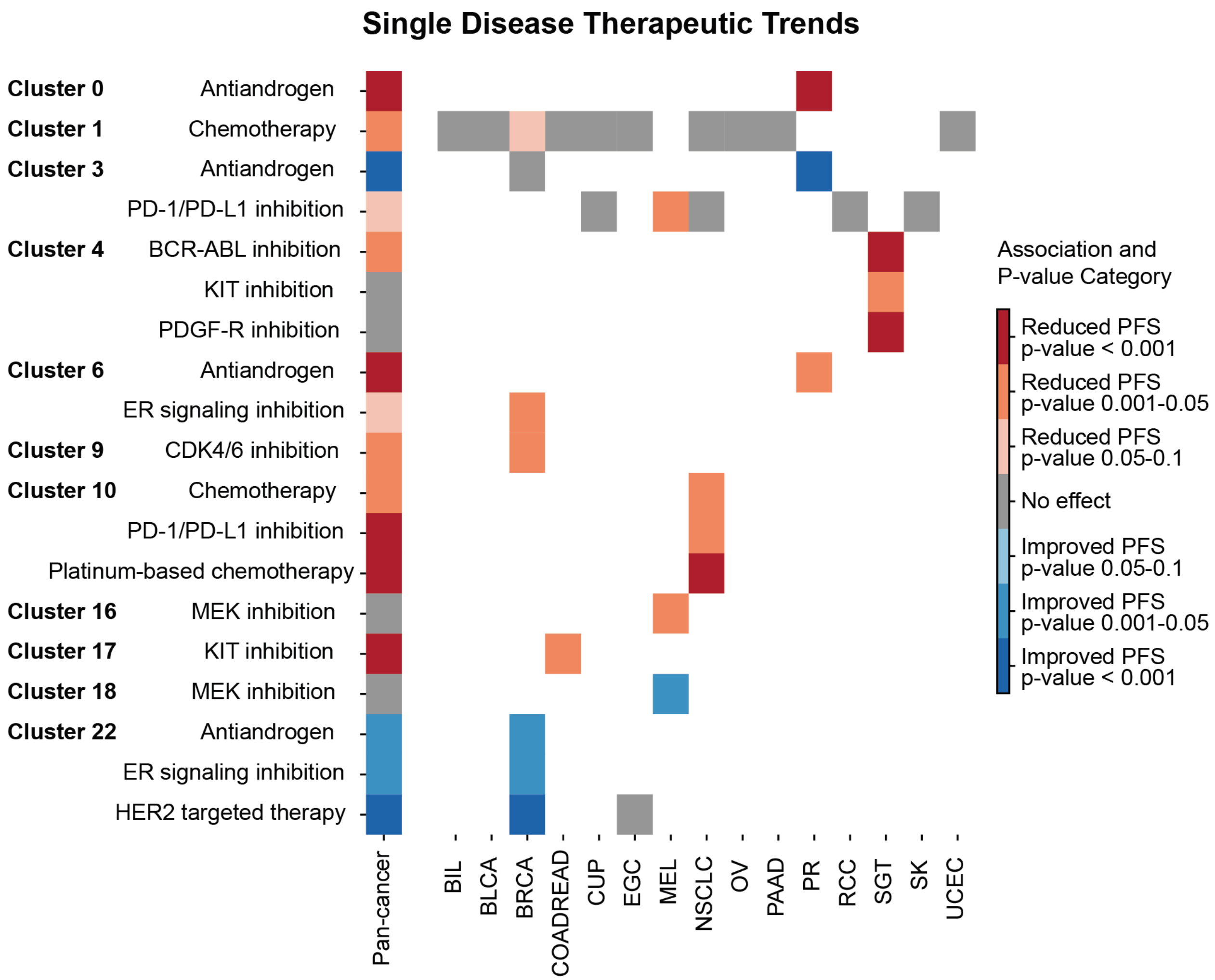


Supplementary Figure 3: Pan-cancer model disease specific clinical trends. Cluster-specific clinical trends with significant or near-significant survival difference in a single cancer type as assessed by log-rank test. Each row is limited to a specific therapy type and considers patients inside vs outside a specific cluster who received that therapy. Each column represents a cohort: the first column is the pan-cancer cohort and the subsequent columns are limited to samples associated with a specific primary diagnosis as listed in the x-axis labels. Specific cancer types included here are the ones with a significant test for at least one cluster-therapy combination included in the plot. The trends shown here are the ones that appear only in a single cancer type. The pan-cancer column indicates whether a trend is still evident in the pan-cancer setting, not that it spans multiple cancer types.

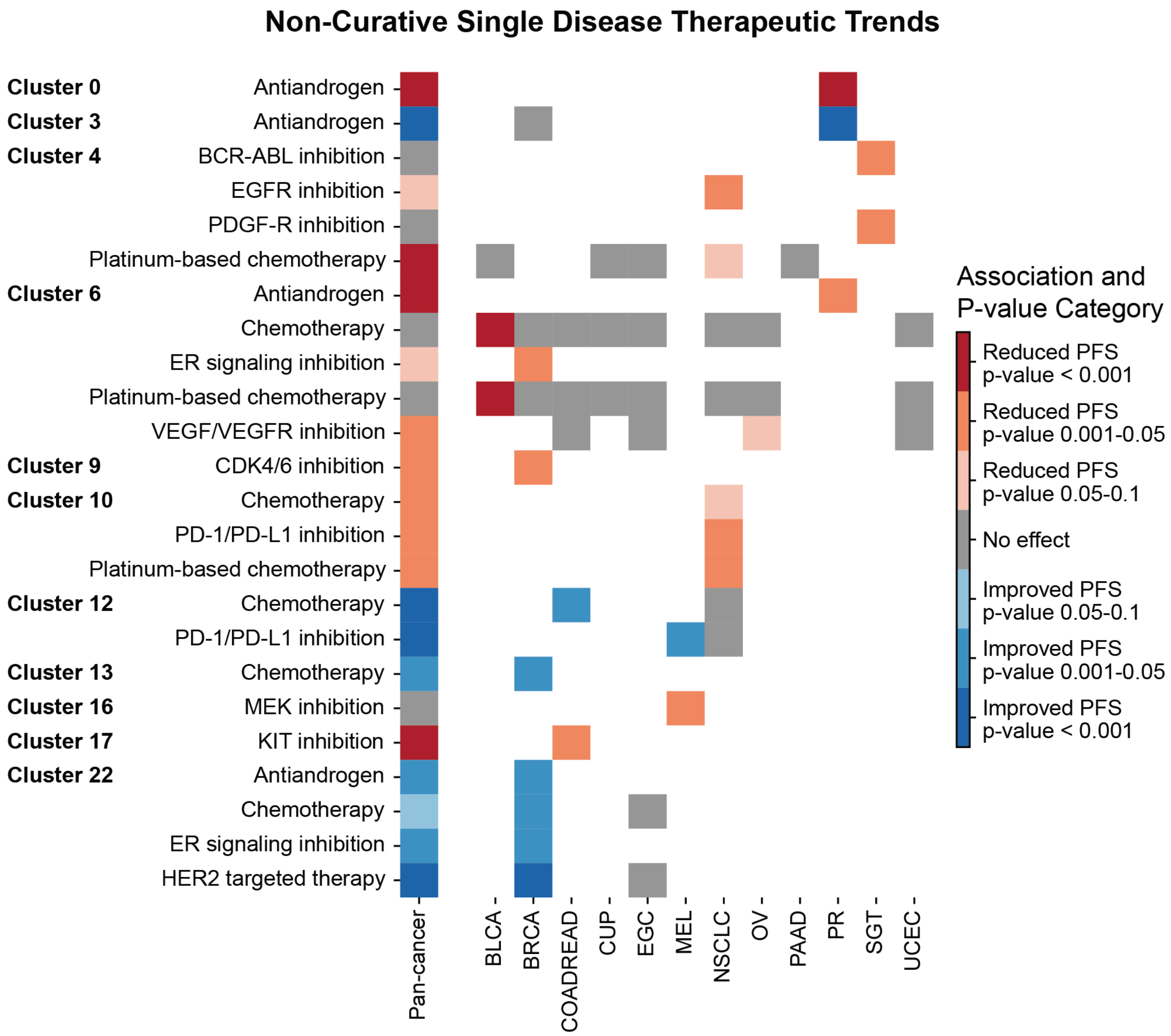


Supplementary Figure 4: Pan-cancer model disease specific clinical trends. Cluster-specific clinical trends with significant or near-significant survival difference in the non-curative setting for a single cancer type as assessed by log-rank test. Each row is limited to a specific therapy type and considers patients inside vs outside a specific cluster who received that therapy with a non-curative intent. Each column represents a cohort: the first column is the pan-cancer cohort and the subsequent columns are limited to samples associated with a specific primary diagnosis as listed in the x-axis labels. Specific cancer types included here are the ones with a significant test for at least one cluster-therapy combination included in the plot. The trends shown are the ones that appear only in a single cancer type. The pan-cancer column indicates whether a trend is still evident in the pan-cancer setting, not that it spans multiple cancer types.

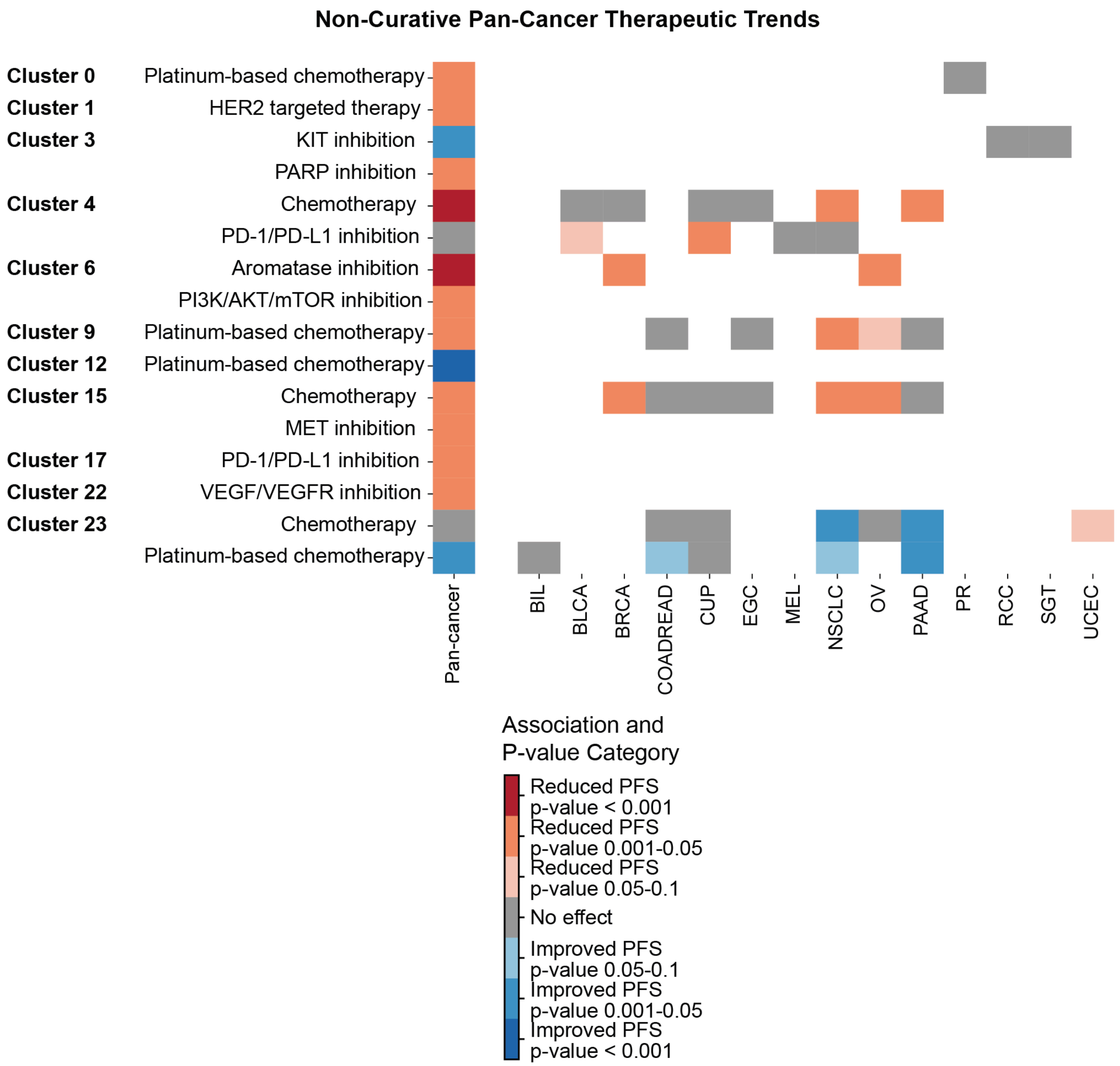


Supplementary Figure 5: Pan-cancer Model Clinical Trends. Cluster-specific clinical trends with significant or near-significant “pan-cancer” survival differences in the non-curative setting as assessed by log-rank test. Each row is limited to a specific therapy type and considers patients inside vs outside a specific cluster who received that therapy with non-curative intent. Each column represents a cohort: the first column is the pan-cancer cohort and the subsequent columns are limited to samples associated with a specific primary diagnosis as listed in the x-axis labels. Specific cancer types included here are the ones with a significant test for at least one cluster-therapy combination included in the plot. The trends shown either appear in more than one cancer type, or are only evaluable/significant in the pan-cancer setting.

**a**

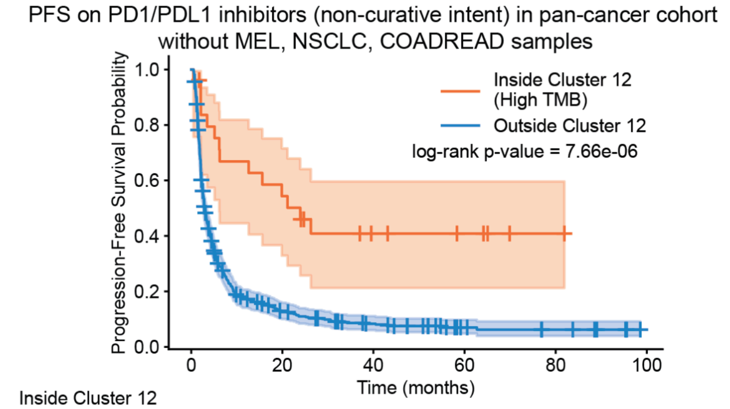


| | 0 | 20 | 40 | 60 | 80 | 100 |
|---|---|---|---|---|---|---|
| **Inside Cluster 12 (High TMB)** | | | | | | |
| At risk | 26 | 13 | 6 | 4 | 1 | 0 |
| Censored | 0 | 2 | 6 | 8 | 11 | 12 |
| Events | 0 | 11 | 14 | 14 | 14 | 14 |
| **Outside Cluster 12** | | | | | | |
| At risk | 529 | 58 | 30 | 10 | 6 | 0 |
| Censored | 0 | 22 | 31 | 47 | 50 | 56 |
| Events | 0 | 449 | 468 | 472 | 473 | 473 |

**b**

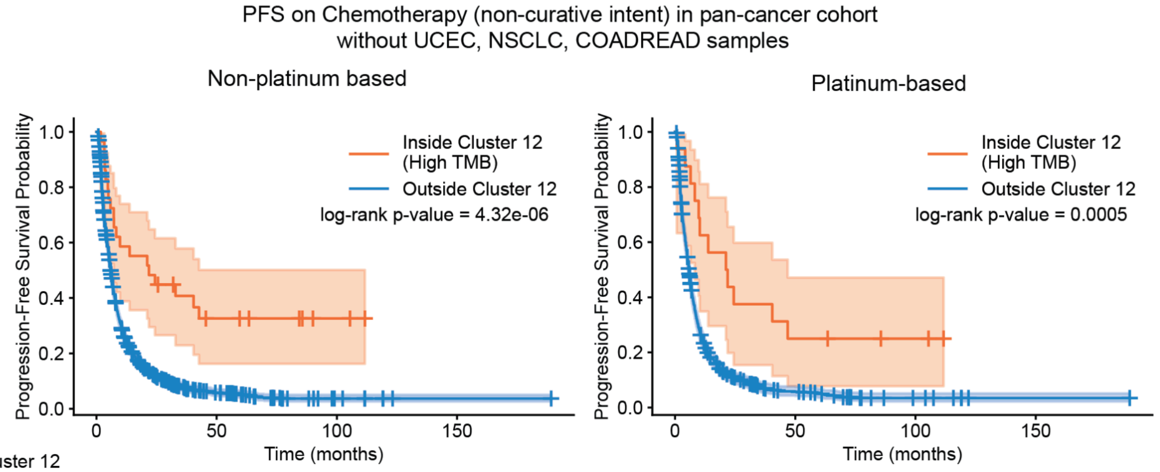


Non-platinum based:

| | 0 | 25 | 50 | 75 | 100 | 125 | 150 | 175 |
|---|---|---|---|---|---|---|---|---|
| **Inside Cluster 12 (High TMB)** | | | | | | | | |
| At risk | 29 | 13 | 7 | 5 | 2 | 0 | 0 | 0 |
| Censored | 0 | 0 | 3 | 5 | 8 | 10 | 10 | 10 |
| Events | 0 | 16 | 19 | 19 | 19 | 19 | 19 | 19 |
| **Outside Cluster 12** | | | | | | | | |
| At risk | 1776 | 175 | 56 | 18 | 7 | 1 | 1 | 1 |
| Censored | 0 | 72 | 116 | 141 | 151 | 157 | 157 | 157 |
| Events | 0 | 1529 | 1604 | 1617 | 1618 | 1618 | 1618 | 1618 |

Platinum-based:

| | 0 | 25 | 50 | 75 | 100 | 125 | 150 | 175 |
|---|---|---|---|---|---|---|---|---|
| **Inside Cluster 12 (High TMB)** | | | | | | | | |
| At risk | 16 | 6 | 4 | 3 | 2 | 0 | 0 | 0 |
| Censored | 0 | 0 | 0 | 1 | 2 | 4 | 4 | 4 |
| Events | 0 | 10 | 12 | 12 | 12 | 12 | 12 | 12 |
| **Outside Cluster 12** | | | | | | | | |
| At risk | 951 | 87 | 34 | 13 | 7 | 1 | 1 | 1 |
| Censored | 0 | 40 | 57 | 68 | 73 | 79 | 79 | 79 |
| Events | 0 | 824 | 860 | 870 | 871 | 871 | 871 | 871 |

Supplementary Figure 6: Pan-cancer model trend in high TMB cluster (cluster 12). a: Improved PFS on PD-1/PD-L1 inhibitors after removing melanoma (MEL), non-small cell lung cancer (NSCLC) and colorectal cancer (COADREAD) samples. b: Improved PFS on chemotherapy (non-platinum based on the left and platinum based on the right) after removing endometrial cancer (UCEC), non-small cell lung cancer (NSCLC) and colorectal cancer (COADREAD) samples.

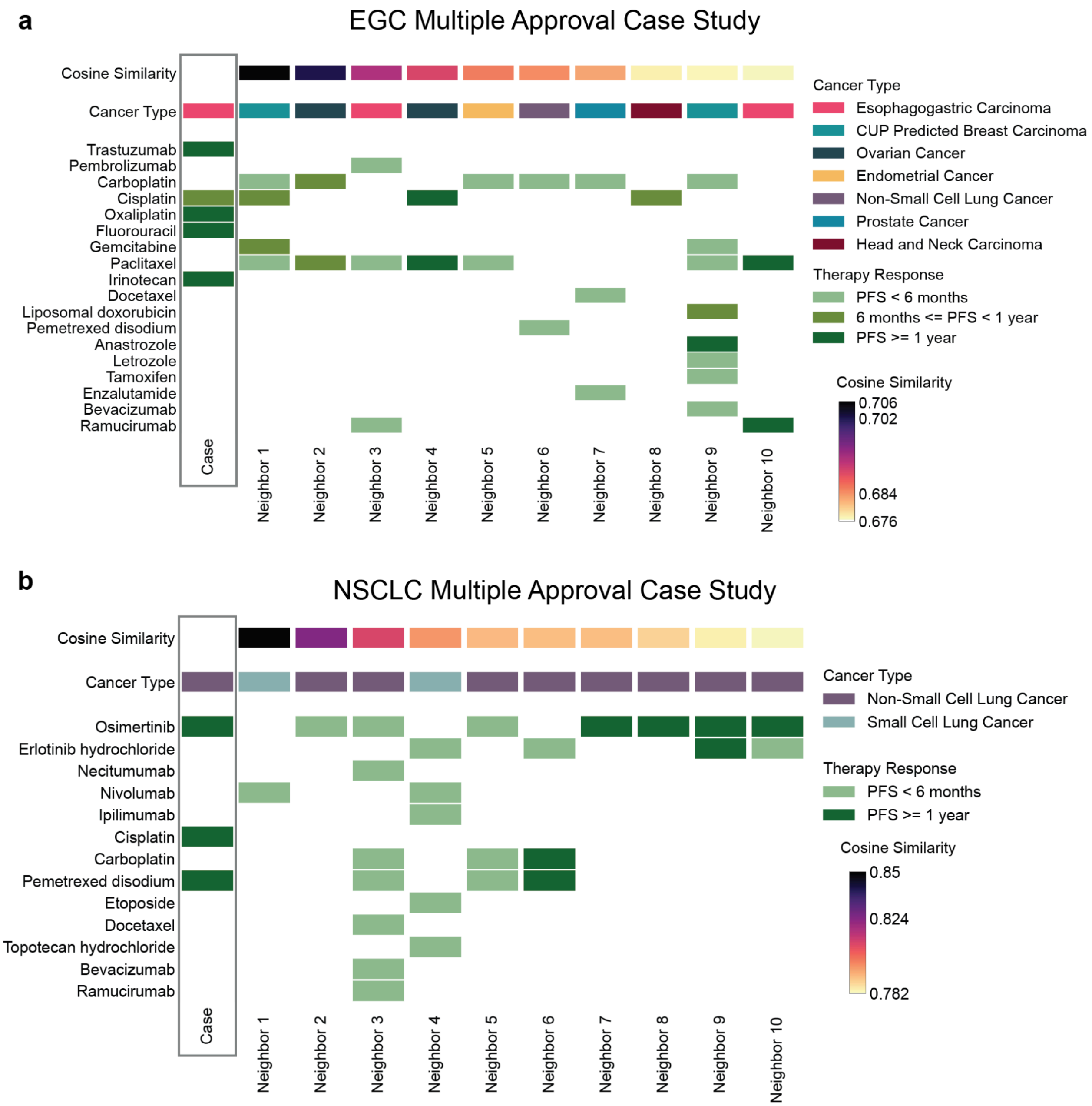


Supplementary Figure 7: Case studies with more than one actionable biomarker. a: Example of an esophagogastric sample with *ERBB2* amplification and high TMB. While current approvals would recommend therapeutic strategies that involve both HER2-directed therapy and PD-1/PD-L1 inhibition, nearest neighbors' low PFS on immunotherapies might guide clinicians away from that option. b: A non-small cell lung cancer case with an *EGFR* mutation and high TMB. Current approvals might recommend both *EGFR* inhibition and PD-1/PD-L1 inhibition, however, the patient neighborhood again suggests that immunotherapies might be ineffective in this case, which is consistent with current management of *EGFR* mutated NSCLC tumors.
